# Treatment outcomes with oral anti-hyperglycaemic therapies in people with diabetes secondary to a pancreatic condition (type 3c diabetes): A population-based cohort study

**DOI:** 10.1101/2024.07.31.24311262

**Authors:** Rhian Hopkins, Katherine G Young, Nicholas J Thomas, Angus G Jones, Andrew T Hattersley, Beverley M Shields, John M Dennis, Andrew P McGovern, the MASTERMIND consortium

## Abstract

**Objectives:** Diabetes secondary to a pancreatic condition (type 3c diabetes) affects 5-10% of people with diabetes, but evidence on the efficacy and tolerability of oral therapies in this group are lacking. We aimed to assess short-term treatment outcomes with oral anti-hyperglycaemic therapies in people with type 3c diabetes.

**Design:** Population-based cohort study.

**Setting:** UK primary care records (Clinical Practice Research Datalink; 2004-2020), linked hospital records.

**Participants:** 7,084 people with a pancreatic condition (acute pancreatitis, chronic pancreatitis, pancreatic cancer, haemochromatosis) preceding diabetes diagnosis (type 3c cohort) initiating an oral glucose-lowering therapy (metformin, sulphonylureas, SGLT2-inhibitors, DPP4-inhibitors, or thiazolidinediones [TZDs]), without concurrent insulin treatment. This cohort was stratified by evidence of pancreatic exocrine insufficiency [PEI] (1,167 with PEI, 5,917 without) and matched to 97,227 type 2 diabetes (T2D) controls.

**Main outcome measures:** 12-month HbA1c change and treatment discontinuation within 6 months, in the type 3c diabetes cohort compared to T2D controls.

**Results:** People with type 3c diabetes had a substantial mean HbA1c reduction with oral therapies in those with PEI (9.4 mmol/mol [95%CI 8.9 to 10.0]) and without (12.2 mmol/mol [12.0 to 12.4]). Compared to T2D controls, people with type 3c diabetes without PEI had a similar mean HbA1c reduction (0.7 mmol/mol [0.4 to 1.0] difference) and similar odds of early treatment discontinuation (Odds ratio [OR] 1.08 [0.98 to 1.19]). In contrast, people with type 3c diabetes and PEI had a lower mean HbA1c response (3.5 mmol/mol [2.9 to 4.1] lesser reduction), and greater discontinuation (OR 2.03 [1.73 to 2.36]). Results were largely consistent across type 3c subtypes and individual drug classes.

**Conclusions:** Oral anti-hyperglycaemic therapies are effective in people with type 3c diabetes, and could provide an important component of glycaemic management. However, the presence of PEI is associated with modestly reduced glycaemic response and reduced tolerability, meaning PEI could identify people that may benefit from closer monitoring after initiating oral therapy.

**What is already known on this topic:**

- Diabetes secondary to a pancreatic condition (type 3c diabetes) is common, affecting 5-10% of people with diabetes in Western populations.
- People with type 3c diabetes are commonly excluded from major diabetes drug trials, meaning there is a lack of management guidelines and evidence on the efficacy and tolerability of oral glucose-lowering therapies in this group.

**What this study adds:**

- Oral glucose-lowering therapies are effective at treating hyperglycaemia in non-insulin treated type 3c diabetes, with largely similar responses to type 2 diabetes observed, and could provide an important component of glycaemic management.
- Pancreatic exocrine insufficiency is associated with modestly reduced glycaemic response and reduced tolerability.

## Introduction

Type 3c diabetes, also known as diabetes of the exocrine pancreas or pancreatogenic diabetes, results from damage to the pancreas and can be caused by a variety of pancreatic conditions. These include acute pancreatitis, chronic pancreatitis, pancreatic cancer, cystic fibrosis, haemochromatosis, and surgical pancreatic resection. [1-7] Type 3c diabetes comprises 5-10% of people with diabetes in Western populations. [8] The adult incidence is higher than that of type 1 diabetes, [9] and may also be increasing. [10] In clinical practice, people with type 3c diabetes are most commonly classified as having type 2 diabetes, but have worse glycaemic control and a greater requirement for insulin. [9, 11]

Guidelines for the management of type 3c diabetes are limited and evidence to inform treatment decisions is inadequate. [8, 12, 13] Treatment of hyperglycaemia in type 3c diabetes is not clearly differentiated from that for type 1 and type 2 diabetes, [6] with current recommendations limited to advising early insulin initiation and avoidance of incretin-based therapies due to their potential association with pancreatitis. [12] There is a lack of studies providing evidence on whether oral diabetes therapies are effective and safe in the management of hyperglycaemia in type 3c diabetes. [6, 7, 14] There are also no dedicated randomized controlled trials of treatments in type 3c diabetes, and people with this type of diabetes are usually excluded from major diabetes drug trials. [2, 8, 15] Due to misclassification of diabetes type and a lack of treatment guidelines, a substantial proportion of people with this type of diabetes may be receiving glucose-lowering therapies that are sub-optimal. [15, 16]

Oral glucose-lowering therapies could potentially be less effective in type 3c diabetes, and their efficacy likely varies by the degree of pancreatic damage. However, treatment response in these patients has not been systematically assessed. Type 3c diabetes is a very heterogenous condition, [7, 17] and therefore the different underlying causes could also potentially result in different treatment responses. In addition to the endocrine dysfunction that results in diabetes, the exocrine functions of the pancreas can be damaged resulting in pancreatic exocrine insufficiency (PEI). [7, 9] This arises when the production of pancreatic enzymes that typically digest food and absorb nutrients is impaired, [6] and is a key feature of this type of diabetes. [12] We hypothesised that PEI could be a marker of the severity of pancreatic damage and could therefore predict efficacy of oral therapies.

We aimed to assess treatment outcomes to oral anti-hyperglycaemic therapies in people with diabetes secondary to a pancreatic condition (type 3c diabetes) and compare them to those with type 2 diabetes.

## Methods

### Data sources

UK population-based primary care data from the Clinical Practice Research Datalink (CPRD), a large database of routinely collected medical records, covering demographics, diagnoses, prescriptions, and test results from over 2,000 primary care practices. [18, 19] CPRD data were linked to national Hospital Episode Statistics (HES), and Index of Multiple Deprivation data (national measure of deprivation).

### Study participants

We extracted primary care records and linked hospital records on all people with a diabetes diagnosis from 2004 to 2020. People with diabetes were identified from the primary care records using clinical codes. We defined diabetes diagnosis dates as the date of the earliest of a diabetes clinical code, prescription for a glucose-lowering medication, or HbA1c ≥48 mmol/mol.

#### Type 3c diabetes

We identified people with a diagnosis of any type of diabetes who had a pancreatic condition prior to or within 30 days of their diabetes diagnosis date (type 3c diabetes cohort). We defined pancreatic conditions as a record of acute pancreatitis, chronic pancreatitis, pancreatic cancer, or haemochromatosis in the primary or secondary care records. We identified and excluded from our type 3c diabetes cohort anyone with surgical pancreatic resection only or cystic fibrosis prior to diabetes diagnosis as surgical pancreatic resection included both partial resection and total pancreatectomy which were difficult to differentiate between, and the management for cystic fibrosis-related diabetes is already well defined. [20]

We stratified our type 3c diabetes cohort by the presence/absence of pancreatic exocrine insufficiency (PEI) in order to evaluate whether this feature impacts treatment response. We defined PEI as a PEI diagnosis code, or a faecal elastase-1 (FE1) test result less than 200 ug/g, [21] or a prescription for pancreatic enzyme replacement therapy (PERT), prior to diabetes diagnosis.

#### Type 2 diabetes

As a comparator group, we identified people with type 2 diabetes (T2D) and no record of a prior pancreatic condition. We classified T2D using an algorithm based on diabetes clinical code counts, glucose-lowering medication prescriptions, and diagnosis age. This classification approach has been validated against the use of genetic and biochemical markers of diabetes type. [22] Other types of diabetes were excluded (Supplementary Figure 1).

### Study cohorts

#### Incident cohorts

Clinical and sociodemographic characteristics, and early insulin and oral therapy initiation, were assessed in incident cohorts of people with type 3c diabetes and T2D at the date of their diabetes diagnosis (Supplementary Figure 1).

#### Treatment outcome cohorts

For both type 3c diabetes and T2D, treatment outcome cohorts were defined as people initiating a major oral glucose-lowering therapy class (metformin, sulphonylureas, thiazolidinediones, DPP4-inhibitors, or SGLT2-inhibitors) for the first time, with the index date set to the date of initiation of that therapy class. GLP-1 receptor agonists were not evaluated as they were initiated by only 258 people with type 3c diabetes. If an individual initiated more than one therapy class over the study period, they were eligible for inclusion in evaluation of each initiated drug class. We excluded people treated with concurrent insulin therapy, those with T2D who developed a pancreatic condition before drug initiation or with a record of PEI at any time, and those with type 3c diabetes with a record of PEI only after drug initiation (Supplementary Figure 2). People with type 3c diabetes were then matched to up to 10 T2D controls initiating the same drug class, without replacement, using a combination of exact matching by sex, ethnicity, deprivation, age (10-year bands), calendar year (5-year bands) and the number of previously initiated anti-hyperglycaemic drug classes (0,1,2+), and nearest-neighbour matching by continuous age and calendar year of drug initiation.

### Outcomes and covariates

In the incident cohort we defined time to first insulin prescription and time to first oral agent prescription from diabetes diagnosis. In the treatment outcome cohort we defined 3 outcomes: HbA1c response, early discontinuation, and weight change. HbA1c response and weight change were defined as the change from baseline 12 months after drug initiation (the closest HbA1c/weight measure to 12 months after initiation within 3-15 months) on unchanged therapy (no addition or cessation of other glucose-lowering medications, and continued prescription of the drug of interest). Early treatment discontinuation was defined as the discontinuation of a therapy within 6 months of initiation, with the availability of at least 3 months follow-up required to confirm discontinuation.

We extracted baseline sociodemographic and clinical features comprising: sex, current age, ethnicity (White, south Asian, Black, Other, Mixed), Index of Multiple Deprivation quintiles, baseline HbA1c (closest value within 6 months before and 7 days after index date), and baseline BMI and weight (closest values within 2 years before and 7 days after index date), number of other glucose-lowering therapies currently being taken, and alcohol consumption (none, within government recommended limits, exceeding recommended limits, clinically harmful). Variables were defined using primary care records with the exception of deprivation.

### Statistical analysis

In the incident cohort, we assessed time to insulin initiation and time to oral agents up to 3 years from diabetes diagnosis by PEI status and by type 3c diabetes subtype, using Kaplan Meier survival curves and unadjusted Cox proportional hazards models, censoring for death, the end of an individual’s GP records or end of the study period.

In the treatment outcome cohort, our exposure of interest was diabetes type: type 3c diabetes with PEI, type 3c diabetes without PEI, or T2D. We evaluated HbA1c response and weight changes using linear regression, adjusted for baseline measures. Discontinuation was evaluated using logistic regression, adjusted for baseline HbA1c. All models were adjusted for the number of other glucose-lowering therapies being taken. To allow comparison of treatment outcomes by drug class, adjusted estimates of HbA1c response and discontinuation were standardised to baseline HbA1c of 73 mmol/mol, and weight change to baseline weight of 93 kg. All outcomes were standardised for 1 other glucose-lowering therapy currently being taken (0 if the drug class was metformin as this is standard first-line treatment). Analysis was conducted overall, stratified by type 3c diabetes subtype, and by drug class (with the exception of weight change as different drug classes are known to have opposite effects on weight change [23]). For all models, T2D controls were weighted as the inverse of the number of controls matched to an individual with type 3c diabetes. People with missing covariate or outcome data were excluded from each analysis.

### Sensitivity analysis

Alcoholism is a common cause of pancreatitis, and also negatively affects medication compliance in chronic conditions. [24] To assess the potential impact of this, we repeated the overall treatment response analysis excluding people with a history of excess alcohol usage. In order to assess if our results were sensitive to our PEI definition, we also repeated overall analysis restricting the PEI subgroup definition to require a PERT prescription in the 6 months prior to drug initiation. To assess our definition of diabetes secondary to acute pancreatitis, where it is likely we will have included some people with T2D unrelated to their prior pancreatic condition, we repeated overall analysis in this subgroup in people with a record of acute pancreatitis within the 5 years prior to diabetes diagnosis.

Further information on study methods is available online (code lists: https://github.com/Exeter-Diabetes/CPRD-Codelists, defining cohorts: https://github.com/Exeter-Diabetes/CPRD-Cohort-scripts/, analysis: https://github.com/Exeter-Diabetes/CPRD-Rhian-T3cTreatment-Scripts). Statistical analysis was performed using R version 4.3.0, and RECORD reporting guidelines were followed [25].

### Patient and Public Involvement

The findings of this study were discussed with an individual with type 3c diabetes, which informed the interpretation of results. There was no patient or public involvement in the study design or analysis.

## Results

### Type 3c diabetes clinical characteristics at diagnosis

We identified 10,318 people with type 3c diabetes (1,518 with PEI and 8,800 without PEI) and 524,084 people with T2D and no prior pancreatic conditions (Table 1).

**Table 1.** Baseline clinical and sociodemographic characteristics at diabetes diagnosis and treatment after diagnosis in the incident cohorts of individuals with type 3c diabetes and type 2 diabetes controls.

|  | Type 2 diabetes | Overall | Type 3c diabetes<br>PEI | No PEI |
| --- | --- | --- | --- | --- |
| n | 524084 | 10318 | 1518 | 8800 |
| **Sex** |  |  |  |  |
| Male | 295514 ( 56.4) | 6071 (58.8) | 965 (63.6) | 5106 (58.0) |
| Female | 228570 ( 43.6) | 4247 (41.2) | 553 (36.4) | 3694 (42.0) |
| **Age, years** |  |  |  |  |
| Median [IQR] | 60.90 [51.00, 70.67] | 60.30 [49.60, 70.71] | 58.38 [48.28, 67.70] | 60.74 [49.82, 71.35] |
| **Ethnicity** |  |  |  |  |
| White | 421553 ( 82.7) | 9265 (90.6) | 1432 (94.7) | 7833 (89.9) |
| South Asian | 51054 ( 10.0) | 536 ( 5.2) | 40 ( 2.6) | 496 ( 5.7) |
| Black | 25492 ( 5.0) | 249 ( 2.4) | 27 ( 1.8) | 222 ( 2.5) |
| Other | 7142 ( 1.4) | 91 ( 0.9) | 7 ( 0.5) | 84 ( 1.0) |
| Mixed | 4670 ( 0.9) | 80 ( 0.8) | 6 ( 0.4) | 74 ( 0.8) |
| **Index of multiple deprivation quintile** |  |  |  |  |
| 1 (least deprived) | 93211 ( 18.0) | 1790 (17.5) | 253 (16.8) | 1537 (17.7) |
| 2 | 97264 ( 18.8) | 1777 (17.4) | 277 (18.4) | 1500 (17.2) |
| 3 | 100360 ( 19.4) | 1954 (19.2) | 279 (18.6) | 1675 (19.3) |
| 4 | 109983 ( 21.2) | 2189 (21.5) | 308 (20.5) | 1881 (21.6) |
| 5 (most deprived) | 117611 ( 22.7) | 2492 (24.4) | 387 (25.7) | 2105 (24.2) |
| **HbA1c, mmol/mol** |  |  |  |  |
| Median [IQR] | 53.00 [48.81, 67.10] | 53.00 [49.00, 72.67] | 54.04 [49.00, 78.44] | 53.00 [49.00, 71.79] |
| **BMI, kg/m2** |  |  |  |  |
| Median [IQR] | 31.60 [27.90, 36.10] | 29.64 [25.40, 34.20] | 24.80 [21.30, 28.83] | 30.30 [26.30, 34.90] |
| **3c subtype** |  |  |  |  |
| Acute pancreatitis only | NA | 5322 (51.6) | 129 ( 8.5) | 5193 (59.0) |
| Chronic pancreatitis | NA | 3623 (35.1) | 1095 (72.1) | 2528 (28.7) |
| Haemochromatosis | NA | 681 ( 6.6) | 3 ( 0.2) | 678 ( 7.7) |
| Pancreatic cancer | NA | 692 ( 6.7) | 291 (19.2) | 401 ( 4.6) |

|  | Type 2<br>diabetes | Overall | Type 3c diabetes |  |
| --- | --- | --- | --- | --- |
|  |  |  | PEI | No PEI |
| Treatment after diagnosis |  |  |  |  |
| Insulin within 1 year, % (95% CI) | 2.0 [1.9,2.0] | 18.4 [17.7,19.2] | 32.8 [30.4,35.2] | 15.9 [15.2,16.7] |
| Insulin within 3 years, % (95% CI) | 3.5 [3.4,3.5] | 23.7 [22.9,24.6] | 43.7 [41.1,46.2] | 20.3 [19.5,21.2] |
| Oral agent within 1 year, % (95% CI) | 49.8 [49.7,49.9] | 51.0 [50.0,51.9] | 48.9 [46.4,51.4] | 51.3 [50.3,52.3] |
| Oral agent within 3 years, % (95% CI) | 64.8 [64.7,65.0] | 65.1 [64.1,66.0] | 60.4 [57.8,62.9] | 65.9 [64.8,66.9] |

People with type 3c diabetes, particularly those with PEI, were more commonly male, of White ethnicity, and had greater deprivation, compared to those with T2D. Those with type 3c diabetes and PEI also had a lower BMI. HbA1c was largely similar between groups. People with type 3c diabetes and PEI most commonly had prior chronic pancreatitis (72.1%), followed by prior pancreatic cancer (19.2%), whereas the majority of those with type 3c diabetes without PEI had prior acute pancreatitis (59.0%).

### Treatment after diagnosis

Early insulin initiation was more common in people diagnosed with type 3c diabetes compared to people with T2D, most marked in those with PEI (PEI: 43.7% insulin treated at 3 years; hazard ratio [HR] 17.27 [95% CI 15.96-18.69], without PEI: 20.3% insulin treated at 3 years; HR 6.79 [95% CI 6.46-7.13] [Table 1, Supplementary Figure 3]). There was also heterogeneity between type 3c diabetes subtypes, with the greatest early insulin initiation in pancreatic cancer (HR 19.48 [95% CI 17.37-21.84]) and chronic pancreatitis subtypes (HR 13.82 [95% CI 13.06-14.62]) (Supplementary Figure 4). The proportion of people initiating oral agents following diabetes diagnosis was largely similar between people with type 3c diabetes and those with T2D, although there was less oral therapy initiation at 3 years in those with PEI (HR 0.93 [95% CI 0.87-1.00]) and those with pancreatic cancer (HR 0.83 [95% CI 0.75-0.92]) (Supplementary Figures 5-6).

### Overall oral therapy outcomes

7,084 people with type 3c diabetes initiating an oral glucose-lowering therapy (1,167 with PEI and 5,917 without) were matched to 97,227 T2D controls (baseline characteristics in Supplementary Table 1).

In the type 3c diabetes treatment outcome cohort, a substantial mean HbA1c response with oral therapy was observed: 12.2 mmol/mol (95% CI 12.0-12.4) in people without PEI and 9.4 mmol/mol (95% CI 8.9-10.0) in people with PEI (Figure 1A). Compared to the T2D controls, HbA1c response was largely similar in those without PEI (0.7 [95% CI 0.4-1.0] mmol/mol lesser mean HbA1c response) but lower in those with PEI (3.5 [95% CI 2.9-4.1] mmol/mol lesser response).

**Figure 1.**
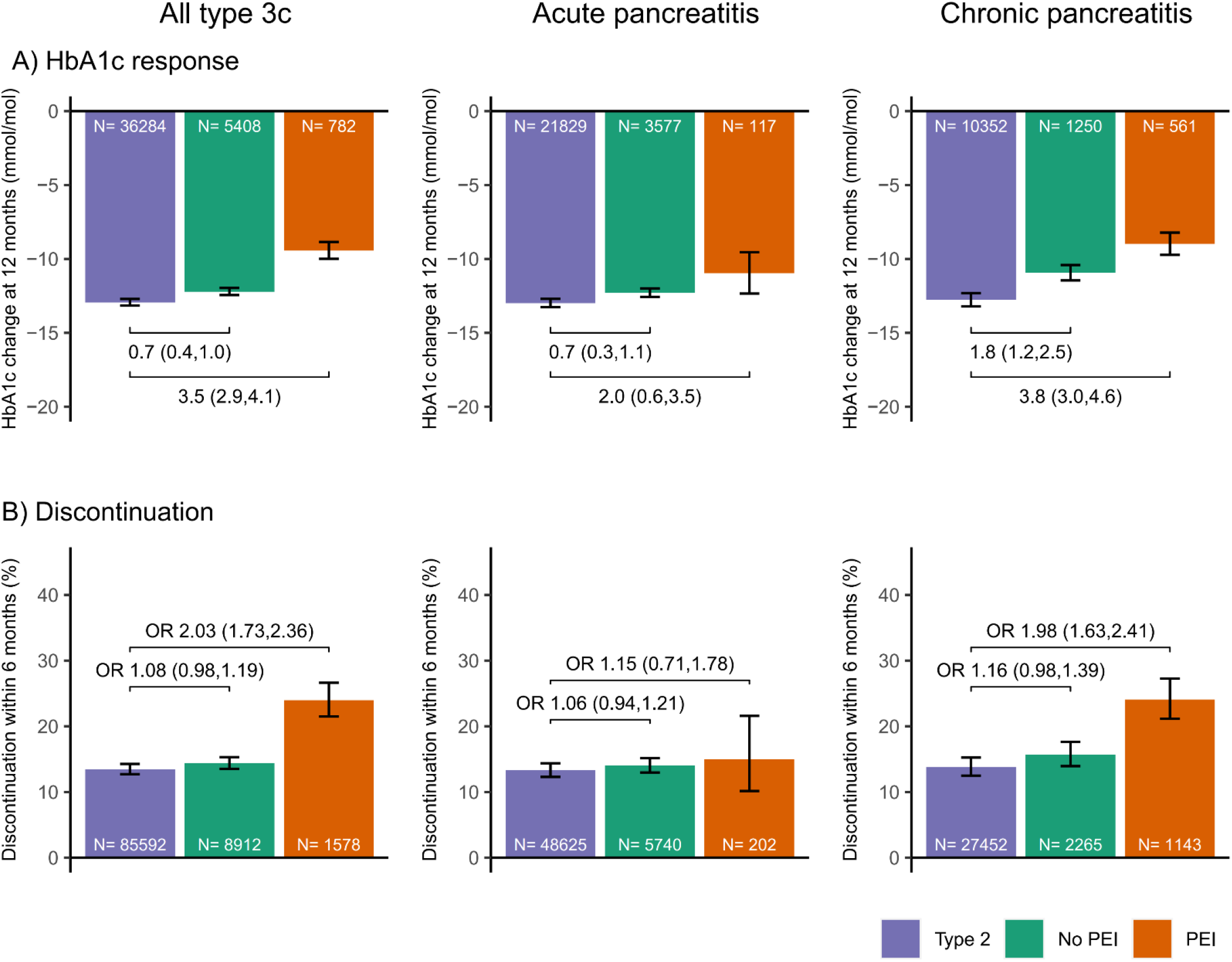
A) Mean HbA1c response and B) Proportion of early treatment discontinuation, in individuals with type 3c diabetes and PEI (orange), type 3c diabetes without PEI (green), and matched type 2 diabetes controls (blue/purple) initiating an oral glucose lowering therapy, for the full type 3c cohort, those with type 3c diabetes following acute pancreatitis, and those with type 3c diabetes following chronic pancreatitis. Contrasts represent estimated differences between groups with 95% confidence intervals for HbA1c change in mmol/mol and odds ratios with 95% confidence intervals for discontinuation. Models were adjusted for baseline HbA1c and number of other glucose-lowering therapies being taken.

Early discontinuation of oral therapy within 6 months was 11.2% in the T2D controls, 12.8% in those with type 3c diabetes without PEI, and 20.0% in those with PEI. Compared to T2D controls, the odds of discontinuation in people with type 3c diabetes were similar in those without PEI (Odds ration [OR] 1.08 [95% CI 0.98-1.19]) but higher in those with PEI (OR 2.03 [95% CI 1.73-2.36]; Figure 1B).

### Oral therapy outcomes by Type 3c diabetes subtype

There was a similar pattern of treatment outcomes when assessing type 3c diabetes subtypes, with a modestly reduced glycaemic response in those with type 3c diabetes for both chronic pancreatitis and acute pancreatitis subtypes compared to T2D controls, most marked in those with PEI (Figure 1A). Discontinuation was higher versus T2D controls in the chronic pancreatitis subgroup with PEI, with no evidence of a difference for other subgroups (Figure 1B). In those with pancreatic cancer and PEI, there was reduced glycaemic response (3.1 [95% CI 1.3-4.9] mmol/mol lesser HbA1c reduction) and increased discontinuation compared to T2D controls (OR 2.60 [95% CI 1.60-4.25]), with no difference in either outcome for those without PEI (Supplementary Figure 7). In the haemochromatosis subgroup, only 4 people had PEI (treatment outcome differences therefore not analysed) and in those without PEI glycaemic response and discontinuation were similar to the T2D controls (Supplementary Figure 8).

### Oral therapy outcomes by drug class

People with type 3c diabetes showed a substantial HbA1c response across all drug classes. Those without PEI had a slightly lesser response versus T2D controls for sulfonylureas only (1.6 [95% CI 0.9-2.4] mmol/mol lesser response; Figure 2A). In those with PEI, mean response was lower compared to T2D controls for metformin and sulphonylureas (both 4.2 mmol/mol lesser response; Figure 2A). There was no evidence of a reduced response on SGLT2-inhibitors in either group.

**Figure 2.**
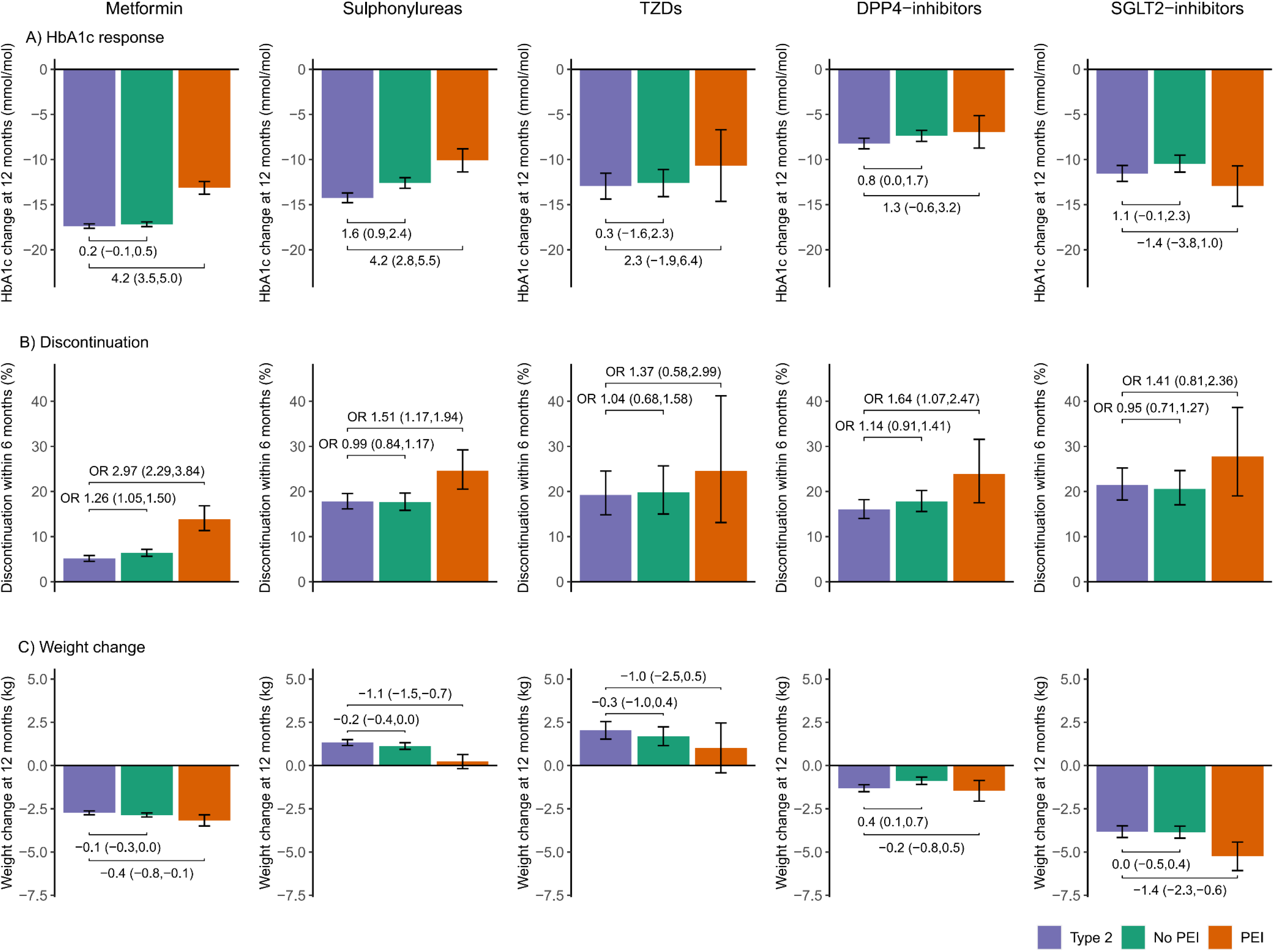
A) Mean HbA1c response, B) Proportion of early treatment discontinuation and C) Mean weight change, in all individuals with type 3c diabetes and PEI (orange), type 3c diabetes without PEI (green), and matched type 2 diabetes controls (blue/purple) initiating an oral glucose lowering therapy, for different classes of oral glucose lowering therapy: Metformin, Sulphonylureas, Thiazolidinediones [TZDs], DPP4-inhibitors, and SGLT2-inhibitors, respectively. Contrasts represent estimated differences between groups with 95% confidence intervals for HbA1c change in mmol/mol and weight chance in kg, and odds ratios with 95% confidence intervals for discontinuation. Models were adjusted for baseline values and number of other glucose-lowering therapies being taken. Numbers in each subgroup are given in Supplementary Table.

Short-term discontinuation in people with type 3c diabetes without PEI was broadly similar to T2D across all drug classes, with the exception of metformin (OR 1.26 [95% CI 1.05-1.50]). In people with type 3c diabetes and PEI discontinuation was greater versus T2D controls across all drug classes, with the most marked increase seen for metformin (OR 2.97 [95% CI 2.29-3.84]; Figure 2B).

Weight change varied by drug class but was largely similar between people with type 3c diabetes and T2D controls (Figure 2C). The exception was for people with type 3c diabetes and PEI, who had greater mean weight loss on SGLT2-inhibitors compared to T2D controls (1.4 [95% CI 0.6-2.3] kg greater weight loss) and lesser mean weight gain on sulphonylureas (1.1 [95% CI 0.7-1.5] kg lesser weight gain). Supplementary tables 2-6 report full numerical treatment outcome results, stratified by both type 3c diabetes subtype and drug class.

### Sensitivity analyses

Treatment outcomes were largely consistent with the primary analysis when 1) including only people whose alcohol consumption was within recommended limits or who did not consume alcohol at drug initiation (Supplementary Figure 9), 2) restricting the cohort of people with type 3c diabetes and PEI to those with a PERT prescription in the 6 months prior to drug initiation (Supplementary Figure 10), and 3) restricting the criteria of diabetes following acute pancreatitis to those with a record of acute pancreatitis within 5 years prior to diabetes diagnosis (Supplementary Figure 11).

## Discussion

In this large UK observational study, we find that people with type 3c diabetes have a substantial HbA1c reduction to five major oral anti-hyperglycaemic drug classes, largely comparable to those with T2D. We also identify clinically relevant heterogeneity within the broad type 3c diabetes phenotype. In particular, PEI, a potential marker of the severity of pancreatic damage, is identified as a novel determinant of treatment outcomes on oral therapy; those with PEI have a modestly reduced glycaemic response and increased risk of early treatment discontinuation compared to people with T2D. In contrast, those without PEI, who make up the majority of our type 3c diabetes cohort, have an only slightly lesser glycaemic response (<1 mmol/mol difference) and similar risk of discontinuation compared to T2D. Findings are broadly consistent across the evaluated drug classes (metformin, sulphonylureas, DPP4-inhibitors, thiazolidinediones, and SGLT2-inhibitors), and suggest that PEI represents a simple clinical marker that may be a more important determinant of treatment outcomes than the underlying pancreatic condition in type 3c diabetes.

Glycaemic response in those with PEI was most attenuated with metformin and sulphonylureas (4.2 mmol/mol lesser response than T2D), although both therapies were still effective. This may be because these medication classes influence the secretion or action of endogenous insulin, processes potentially more adversely affected in type 3c diabetes than T2D, particularly in those with PEI where pancreatic damage may be more severe. A reduced response was not seen for SGLT2-inhibitors, which work independently of endogenous insulin. [15] Metformin discontinuation was the most different between type 3c diabetes and T2D with 3-fold higher odds of discontinuation in those with type 3c diabetes and PEI compared to T2D. This may relate to the known gastro-intestinal side effects of metformin, given PEI causes similar symptoms, and could mean the therapy may not be well-tolerated in some people with type 3c diabetes. [15, 26] The impact of oral medications on weight change was mostly similar in type 3c diabetes and T2D, although small differences for sulphonylureas and SGLT2-inhibitors in those with PEI were observed these are unlikely to be clinically significant.

As previously observed in smaller datasets, we observed greater early insulin initiation in people with type 3c diabetes compared to T2D, [9, 16, 27, 28] particularly in people with chronic pancreatitis or pancreatic cancer. This may be due to more impaired endogenous insulin production as a result of pancreatic damage, [2, 7] but could also reflect the tendency of clinicians to prescribe insulin when type 3c diabetes is suspected given the uncertain effectiveness of oral medications in this patient group.

### Strengths and limitations

To our knowledge, this study is the first to assess short-term outcomes to oral therapies in type 3c diabetes, and whether outcomes vary by PEI status, drug class, and underlying pancreatic condition. A strength of this study is the large size of the dataset enabling analysis of a much larger cohort of people with type 3c diabetes than many defined using other observational datasets. [9, 16, 27-29] However, we acknowledge that when stratifying by subtype and drug class, low numbers still limit these analyses. We were unable to include GLP1 receptor agonists in our study or to assess responses to oral therapies in people concurrently treated with insulin due to low numbers. The lack of a standardised approach to identify type 3c diabetes in routine clinical data is a limitation, and many people with type 3c diabetes will have been labelled with T2D in practice. [9] In the absence of universally accepted diagnostic criteria, we adopted a similar pragmatic approach to previous observational studies of type 3c diabetes and classified people based on prior records of a pancreatic condition. [9, 16, 27, 29] As a result, there will likely be people in our type 3c diabetes cohort who have T2D that may have occurred regardless of their prior pancreatic condition, but this is difficult to determine in our dataset, or indeed in clinical practice. This issue is potentially most likely to occur in our acute pancreatitis subgroup, and so we performed a sensitivity analysis restricting our type 3c diabetes definition to acute pancreatitis close to the date of diabetes diagnosis, which reassuringly showed results consistent with the primary analysis. CPRD data are collected as part of routine care and is widely representative of the general population with many ethnic groups represented. [18] A key strength of this study was the use of both primary care records and hospital admission data to identify records of prior pancreatic conditions in order to identify type 3c diabetes phenotypes as robustly as possible.

### Interpretation

People with type 3c diabetes have commonly been excluded from major diabetes drug trials, [2, 8, 15] and no previous studies have assessed whether and which oral glucose-lowering therapies are effective in treating this group. We have demonstrated that oral glucose-lowering therapies are effective in non-insulin treated people with type 3c diabetes, and for the majority of this group there is no meaningful difference in glycaemic response or tolerability to those with T2D. This suggests that oral therapies could provide an important component of glycaemic management in this group. While oral glucose-lowering therapies remained effective in those with PEI, there was a modest reduction in efficacy and likely reduction in tolerability for most drug classes, in comparison to T2D, suggesting people with type 3c diabetes and PEI may benefit from closer monitoring when making a shared decision to initiate oral diabetes treatment. Future studies should evaluate other patient important outcomes, including the safety of these therapies in people with type 3c diabetes, and address concerns surrounding drug-specific side-effects, for example the risk of hypoglycaemia with sulphonylureas, [15] and the potential association of DPP4-inhibitors and increased risk of pancreatitis. [12]

## Conclusion

In this large UK population-based study, we observed a substantial HbA1c response to five major oral anti-hyperglycaemic therapies in people with type 3c diabetes. By demonstrating that these agents are effective at treating hyperglycaemia in people with type 3c diabetes, our findings suggest oral therapy could provide an important component of glycaemic management. However, PEI was associated with modestly reduced glycaemic response and likely reduced tolerability, and therefore people with PEI may benefit from closer monitoring after initiating oral therapy.

## Data Availability

No additional data are available from the authors although CPRD data are available by application to CPRD Independent Scientific Advisory Committee.

## Acknowledgments

This article is based on data from the Clinical Practice Research Datalink obtained under licence from the UK Medicines and Healthcare products Regulatory Agency. CPRD data is provided by patients and collected by the NHS as part of their care and support. Approval for CPRD data access and the study protocol was granted by the CPRD Independent Scientific Advisory Committee (eRAP protocol numbers: 22_002000). This study was supported by the National Institute for Health and Care Research Exeter Biomedical Research Centre. The views expressed are those of the authors and not necessarily those of the NHS, NIHR or the Department of Health and Social Care. We acknowledge the work of the SPOCC Programme team (https://blogs.exeter.ac.uk/spocc/) and collaborators in the development of code lists for the identification of pancreatic cancer and haemochromatosis phenotypes.

## Funding

This research was funded by the Medical Research Council (UK) (MR/N00633X/1). The funder had no role in any part of the study or in any decision about publication. RH and KGY are supported by Research England’s Expanding Excellence in England (E3) fund. BMS and ATH are supported by the NIHR Exeter Clinical Research Facility. JMD is supported by a Wellcome Trust Early Career award (227070/Z/23/Z).

## Competing interests

APM received prior research funding from Eli Lilly and Company, Pfizer, and AstraZeneca outside of the submitted work. All other authors declare no other relationships or activities that could appear to have influenced the submitted work.

## Author contributions

The study concept and design were developed by APM, JMD, and RH. KGY and RH prepared the data for analysis. RH undertook the analysis, with support from JMD, BMS, and APM. APM, AGJ, NJT, and ATH provided clinical insight. All authors provided support for the interpretation of results, critically revised the manuscript, and saw and approved the final article. APM and JMD attest that all listed authors meet authorship criteria, that no others meeting the criteria have been omitted. APM and JMD are responsible for the decision to submit for publication, and are the guarantors of this work and, as such, had full access to all the data in the study and takes responsibility for the integrity of the data and the accuracy of the data analysis.

## Transparency statement

The lead author affirms that this manuscript is an honest, accurate, and transparent account of the study being reported; that no important aspects of the study have been omitted; and that any discrepancies from the study as planned have been explained.

## Prior presentation

Oral presentation of preliminary results from this study have been given at the Diabetes UK Professional Conference 2024 and the European Association for the Study of Diabetes (EASD) Annual Meeting 2023.

## Ethics/ data approval

The study protocol was approved by the CPRD Independent Scientific Advisory Committee (eRAP protocol numbers: 22_002000).

## Rights Retention

For the purpose of open access, the author has applied a Creative Commons Attribution (CC BY) licence to any Author Accepted Manuscript version arising from this submission.

## References

1. Andersen DK, Korc M, Petersen GM, et al. Diabetes, Pancreatogenic Diabetes, and Pancreatic Cancer. Diabetes. 2017;66(5):1103–10.

2. Wynne K, Devereaux B, Dornhorst A. Diabetes of the exocrine pancreas. J Gastroenterol Hepatol. 2019;34(2):346–54.

3. Hart PA, Bradley D, Conwell DL, et al. Diabetes following acute pancreatitis. Lancet Gastroenterol Hepatol. 2021;6(8):668–75.

4. Ewald N, Bretzel RG. Diabetes mellitus secondary to pancreatic diseases (Type 3c)--are we neglecting an important disease? Eur J Intern Med. 2013;24(3):203–6.

5. Ewald N, Kaufmann C, Raspe A, et al. Prevalence of diabetes mellitus secondary to pancreatic diseases (type 3c). Diabetes Metab Res Rev. 2012;28(4):338–42.

6. Vonderau JS, Desai CS. Type 3c: Understanding pancreatogenic diabetes. JAAPA. 2022;35(11):20–4.

7. Hart PA, Bellin MD, Andersen DK, et al. Type 3c (pancreatogenic) diabetes mellitus secondary to chronic pancreatitis and pancreatic cancer. Lancet Gastroenterol Hepatol. 2016;1(3):226–37.

8. Cui Y, Andersen DK. Pancreatogenic diabetes: special considerations for management. Pancreatology. 2011;11(3):279–94.

9. Woodmansey C, McGovern AP, McCullough KA, et al. Incidence, Demographics, and Clinical Characteristics of Diabetes of the Exocrine Pancreas (Type 3c): A Retrospective Cohort Study. Diabetes Care. 2017;40(11):1486–93.

10. Cho J, Petrov MS. Pancreatitis, Pancreatic Cancer, and Their Metabolic Sequelae: Projected Burden to 2050. Clin Transl Gastroenterol. 2020;11(11):e00251.

11. Olesen SS, Svane HML, Nicolaisen SK, et al. Clinical and biochemical characteristics of postpancreatitis diabetes mellitus: A cross-sectional study from the Danish nationwide DD2 cohort. J Diabetes. 2021;13(12):960–74.

12. American Diabetes Association Professional Practice C. 2. Diagnosis and Classification of Diabetes: Standards of Care in Diabetes-2024. Diabetes Care. 2024;47(Suppl 1):S20–S42.

13. Makuc J. Management of pancreatogenic diabetes: challenges and solutions. Diabetes Metab Syndr Obes. 2016;9:311–5.

14. Petrov MS. DIAGNOSIS OF ENDOCRINE DISEASE: Post-pancreatitis diabetes mellitus: prime time for secondary disease. Eur J Endocrinol. 2021;184(4):R137–R49.

15. Goodarzi MO, Petrov MS. Diabetes of the Exocrine Pancreas: Implications for Pharmacological Management. Drugs. 2023;83(12):1077–90.

16. Viggers R, Jensen MH, Laursen HVB, et al. Glucose-Lowering Therapy in Patients With Postpancreatitis Diabetes Mellitus: A Nationwide Population-Based Cohort Study. Diabetes Care. 2021;44(9):2045–52.

17. Olesen SS, Toledo FGS, Hart PA. The spectrum of diabetes in acute and chronic pancreatitis. Curr Opin Gastroenterol. 2022;38(5):509–15.

18. Wolf A, Dedman D, Campbell J, et al. Data resource profile: Clinical Practice Research Datalink (CPRD) Aurum. Int J Epidemiol. 2019;48(6):1740–g.

19. Clinical Practice Research Datalink. CPRD Data [online]. https://cprd.com/data x(accessed 19 June 2024)

20. Moran A, Brunzell C, Cohen RC, et al. Clinical care guidelines for cystic fibrosis-related diabetes: a position statement of the American Diabetes Association and a clinical practice guideline of the Cystic Fibrosis Foundation, endorsed by the Pediatric Endocrine Society. Diabetes Care. 2010;33(12):2697–708.

21. Phillips ME, Hopper AD, Leeds JS, et al. Consensus for the management of pancreatic exocrine insufficiency: UK practical guidelines. BMJ Open Gastroenterol. 2021;8(1).

22. Thomas NJ, McGovern A, Young KG, et al. Identifying type 1 and 2 diabetes in research datasets where classification biomarkers are unavailable: assessing the accuracy of published approaches. J Clin Epidemiol. 2022;153:34–44.

23. Davies MJ, Aroda VR, Collins BS, et al. Management of Hyperglycemia in Type 2 Diabetes, 2022. A Consensus Report by the American Diabetes Association (ADA) and the European Association for the Study of Diabetes (EASD). Diabetes Care. 2022;45(11):2753–86.

24. Grodensky CA, Golin CE, Ochtera RD, et al. Systematic review: effect of alcohol intake on adherence to outpatient medication regimens for chronic diseases. J Stud Alcohol Drugs. 2012;73(6):899–910.

25. Benchimol EI, Smeeth L, Guttmann A, et al. The REporting of studies Conducted using Observational Routinely-collected health Data (RECORD) statement. PLoS Med. 2015;12(10):e1001885.

26. Rickels MR, Bellin M, Toledo FG, et al. Detection, evaluation and treatment of diabetes mellitus in chronic pancreatitis: recommendations from PancreasFest 2012. Pancreatology. 2013;13(4):336–42.

27. Lee N, Park SJ, Kang D, et al. Characteristics and Clinical Course of Diabetes of the Exocrine Pancreas: A Nationwide Population-Based Cohort Study. Diabetes Care. 2022;45(5):1141–50.

28. Dugic A, Hagstrom H, Dahlman I, et al. Post-pancreatitis diabetes mellitus is common in chronic pancreatitis and is associated with adverse outcomes. United European Gastroenterol J. 2023;11(1):79–91.

29. Cho J, Scragg R, Pandol SJ, et al. Antidiabetic Medications and Mortality Risk in Individuals With Pancreatic Cancer-Related Diabetes and Postpancreatitis Diabetes: A Nationwide Cohort Study. Diabetes Care. 2019;42(9):1675–83.

