## Supplementary Material for "Treatment outcomes with oral anti-hyperglycaemic therapies in people with diabetes secondary to a pancreatic condition (type 3c diabetes): A population-based cohort study"

**Table of Contents**


| Page | |
|---|---|
| **2** | Supplementary Figure 1. Flow diagram of inclusion/exclusion criteria for the incident cohort of individuals with type 3c and type 2 diabetes. |
| **3** | Supplementary Figure 2. Flow diagram of inclusion/exclusion criteria for the treatment response cohort of individuals with type 3c and type 2 diabetes. |
| **4** | Supplementary Figure 3. Kaplan-Meier cumulative incidence curve of time to initiation of insulin within 3 years of diabetes diagnosis for individuals with type 3c diabetes and PEI prior to diabetes diagnosis, type 3c diabetes without PEI prior to diagnosis, and individuals with type 2 diabetes. |
| **5** | Supplementary Figure 4. Kaplan-Meier cumulative incidence curve of time to initiation of insulin within 3 years of diabetes diagnosis for individuals with type 3c diabetes, stratified by subtype, and individuals with type 2 diabetes. |
| **6** | Supplementary Figure 5. Kaplan-Meier cumulative incidence curve of time to initiation of oral glucose-lowering therapy within 3 years of diabetes diagnosis for individuals with type 3c diabetes and PEI prior to diabetes diagnosis, type 3c diabetes without PEI prior to diagnosis, and individuals with type 2 diabetes. |
| **7** | Supplementary Figure 6. Kaplan-Meier cumulative incidence curve of time to initiation of oral glucose-lowering therapy within 3 years of diabetes diagnosis for individuals with type 3c diabetes, stratified by subtype, and individuals with type 2 diabetes. |
| **8-9** | Supplementary Table 1. Baseline characteristics of matched cohort of individuals with type 3c diabetes and type 2 controls initiating a major glucose-lowering therapy class (metformin, sulphonylureas, thiazolidinediones [TZDs], SGLT2-inhibitors, DPP4-inhibitors). |
| **10** | Supplementary Figure 7. A) Mean HbA1c response and B) Proportion of early treatment discontinuation, in individuals with type 3c diabetes following pancreatic cancer with PEI (orange) and without PEI (green), and matched type 2 controls (blue/purple) initiating an oral glucose lowering therapy. |
| **11** | Supplementary Figure 8. A) Mean HbA1c response and B) Proportion of early treatment discontinuation, in individuals with type 3c diabetes following haemochromatosis without PEI (green), and matched type 2 controls (blue/purple) initiating an oral glucose lowering therapy. |
| **12** | Supplementary table 2. Treatment response outcomes by drug class in all individuals with type 3c diabetes and type 2 controls. |
| **13** | Supplementary table 3. Treatment response outcomes by drug class in individuals with type 3c diabetes following acute pancreatitis and type 2 controls. |
| **14** | Supplementary table 4. Treatment response outcomes by drug class in individuals with type 3c diabetes following chronic pancreatitis and type 2 controls. |
| **15** | Supplementary table 5. Treatment response outcomes by drug class in individuals with type 3c diabetes following pancreatic cancer and type 2 controls. |
| **16** | Supplementary table 6. Treatment response outcomes by drug class in individuals with type 3c diabetes following haemochromatosis and type 2 controls. |
| **17** | Supplementary Figure 9. Sensitivity analysis restricted to individuals whose alcohol consumption was within recommended limits or who did not consume alcohol. |
| **18** | Supplementary Figure 10. Sensitivity analysis restricting the criteria of the PEI subgroup to those with a PERT prescription within the 6 months prior to drug initiation. |
| **19** | Supplementary Figure 11. Sensitivity analysis restricting the criteria of diabetes following acute pancreatitis to those with a record of acute pancreatitis within the 5 years prior to diabetes diagnosis. |

**Supplementary Figure 1.** Flow diagram of inclusion/exclusion criteria for the incident cohort of individuals with type 3c and type 2 diabetes.

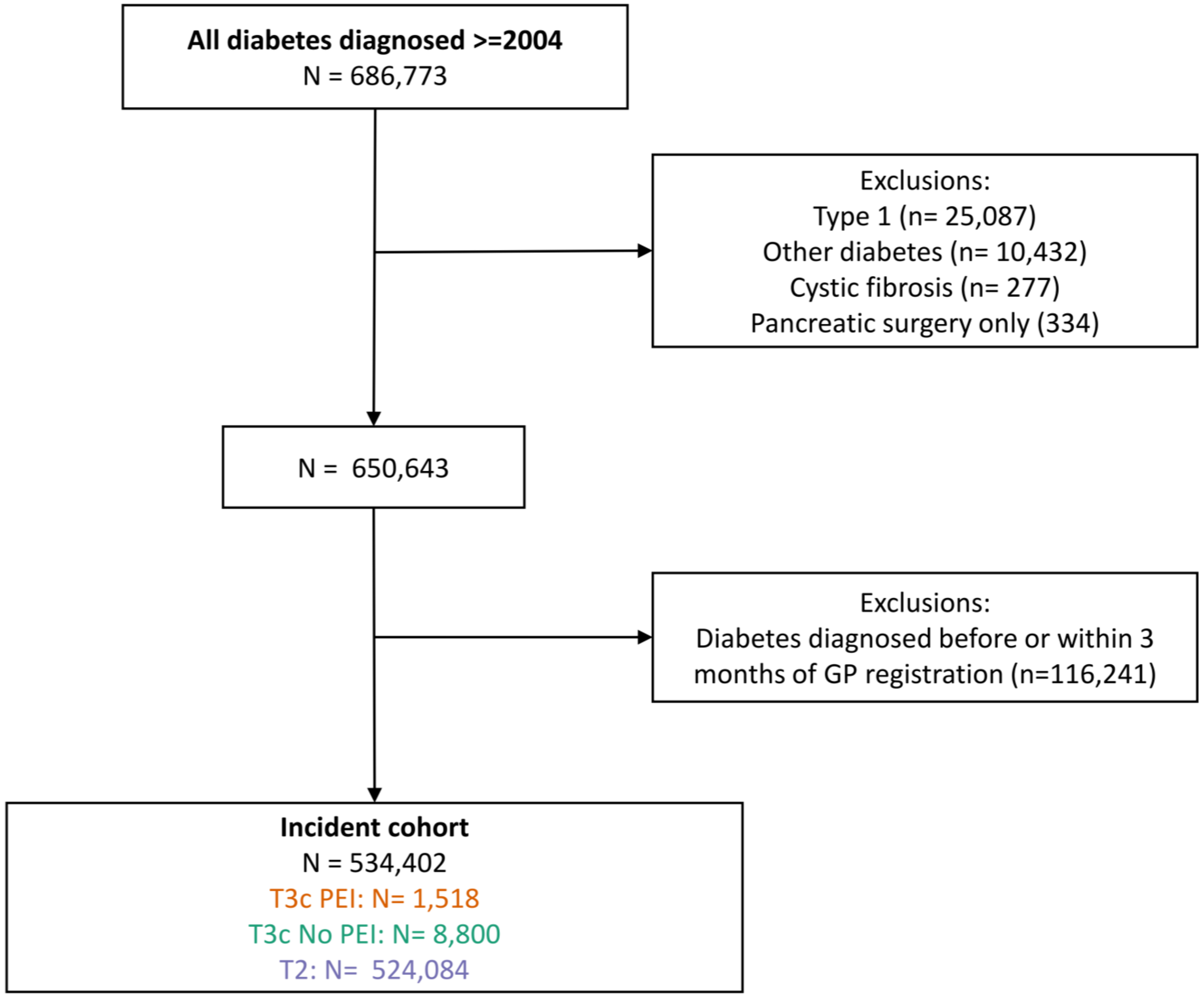

**Supplementary Figure 2.** Flow diagram of inclusion/exclusion criteria for the treatment response cohort of individuals with type 3c and type 2 diabetes.

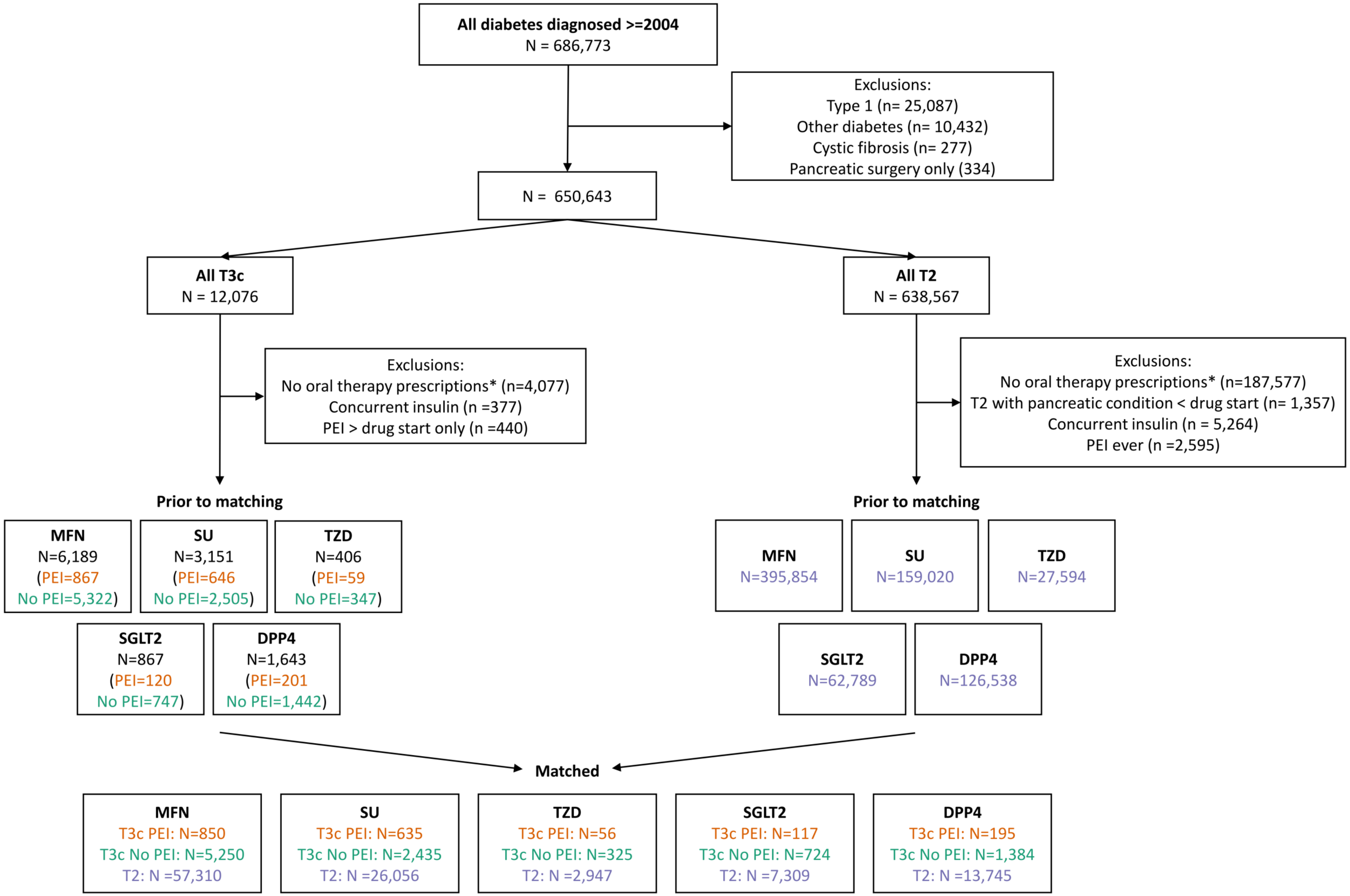

**Supplementary Figure 3.** Kaplan-Meier cumulative incidence curve of time to initiation of insulin within 3 years of diabetes diagnosis for individuals with type 3c diabetes and PEI prior to diabetes diagnosis, type 3c diabetes without PEI prior to diagnosis, and individuals with type 2 diabetes.

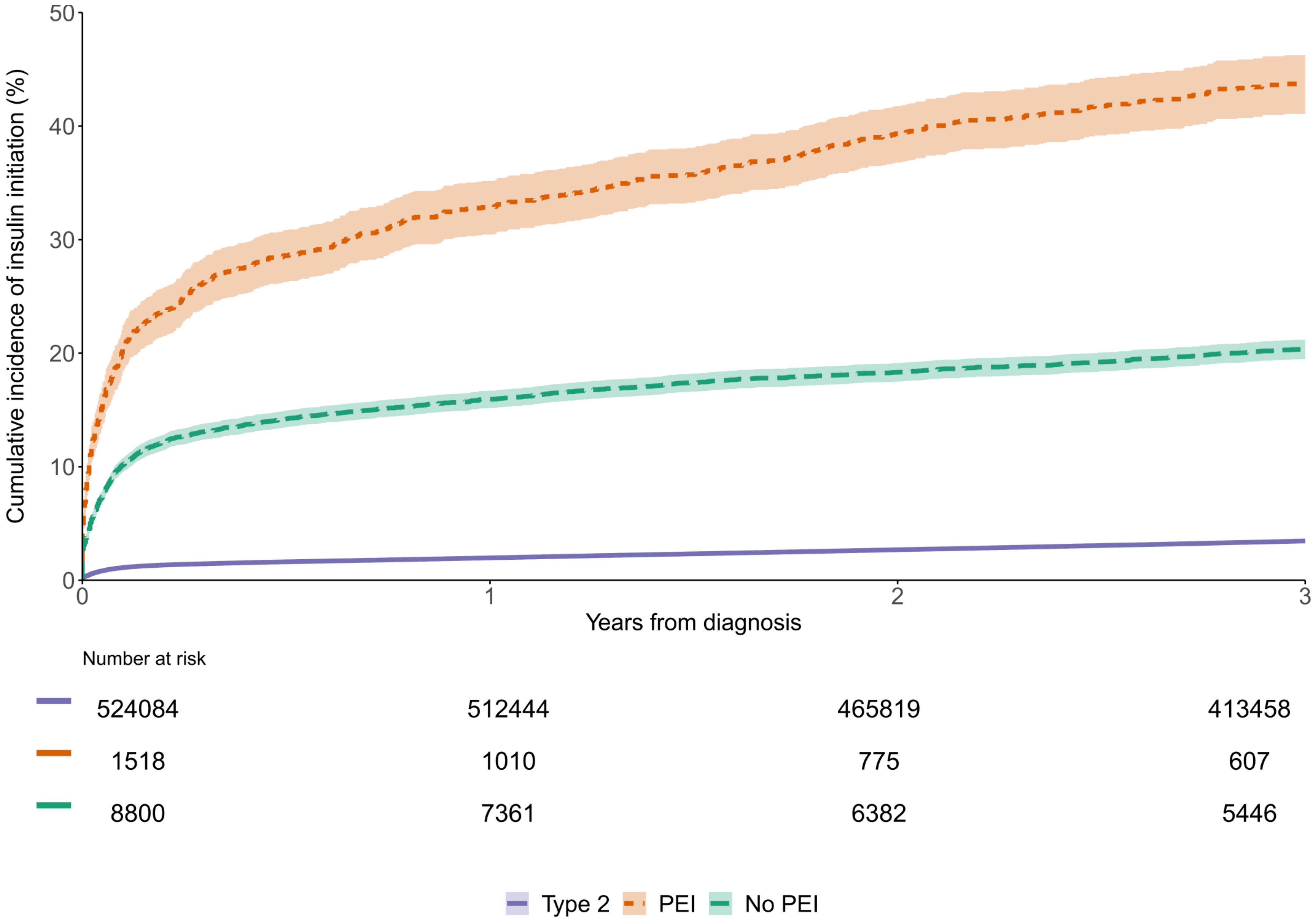

**Supplementary Figure 4.** Kaplan-Meier cumulative incidence curve of time to initiation of insulin within 3 years of diabetes diagnosis for individuals with type 3c diabetes, stratified by subtype, and individuals with type 2 diabetes.

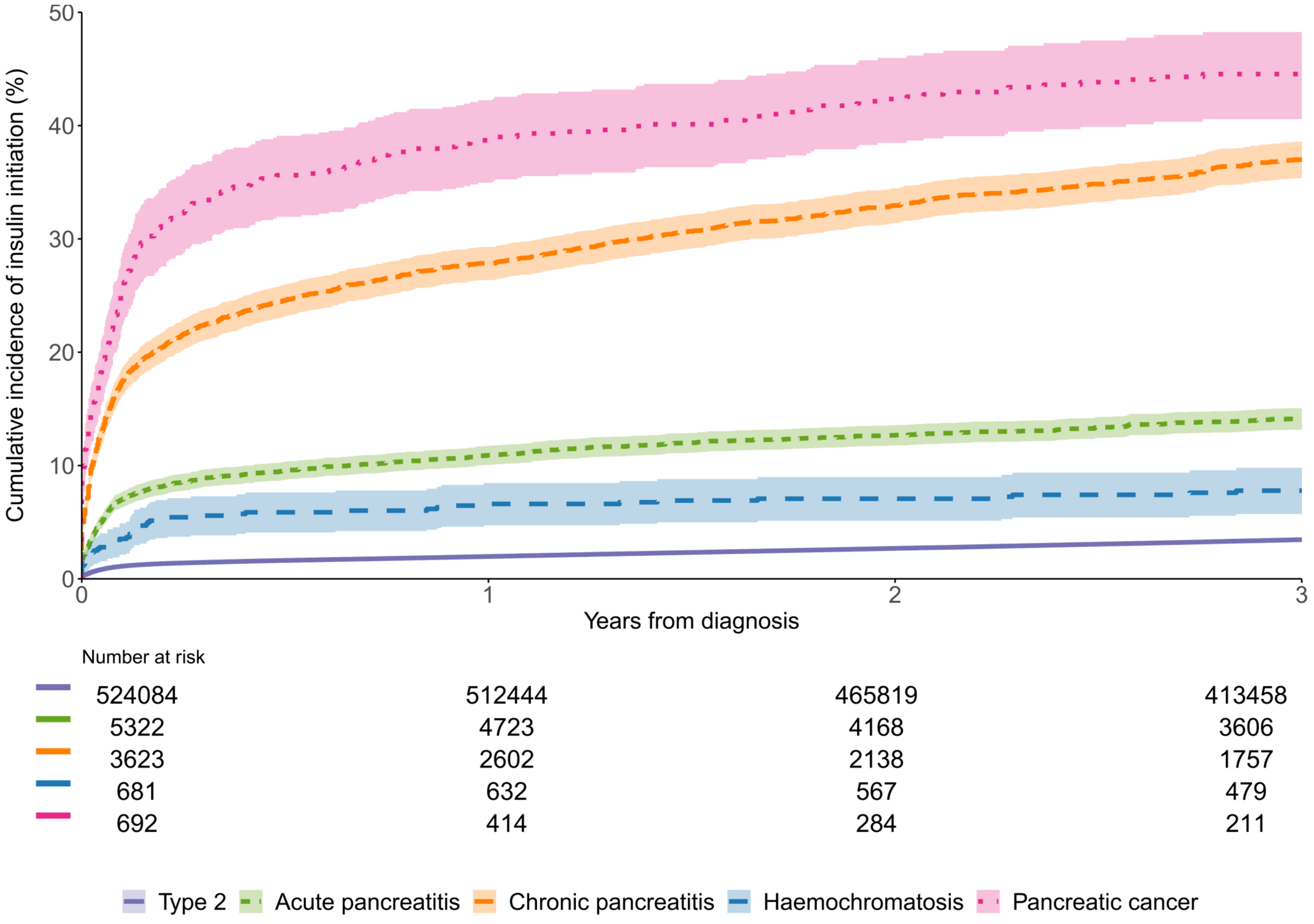

**Supplementary Figure 5.** Kaplan-Meier cumulative incidence curve of time to initiation of oral glucose-lowering therapy within 3 years of diabetes diagnosis for individuals with type 3c diabetes and PEI prior to diabetes diagnosis, type 3c diabetes without PEI prior to diagnosis, and individuals with type 2 diabetes.

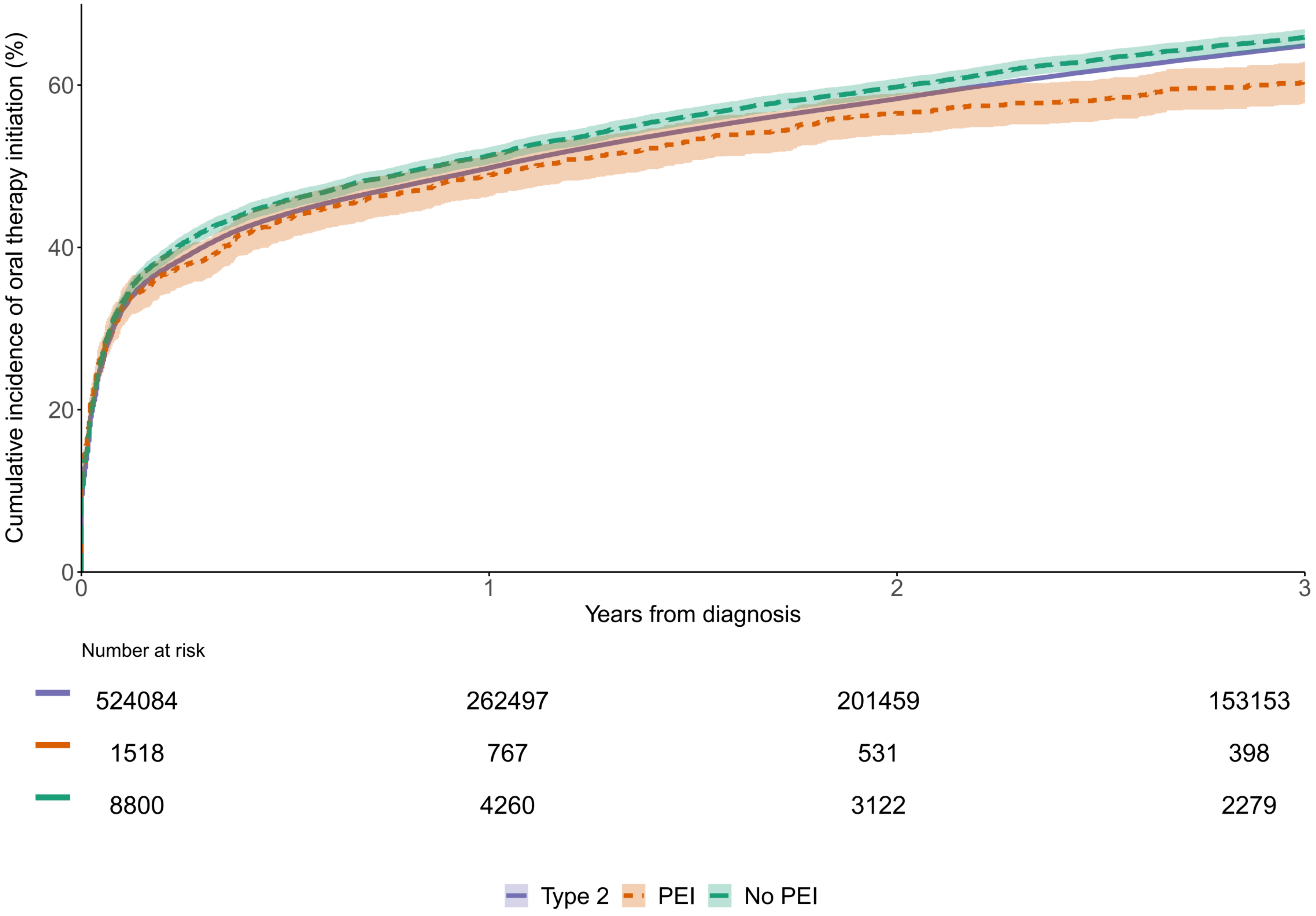

**Supplementary Figure 6.** Kaplan-Meier cumulative incidence curve of time to initiation of oral glucose-lowering therapy within 3 years of diabetes diagnosis for individuals with type 3c diabetes, stratified by subtype, and individuals with type 2 diabetes.

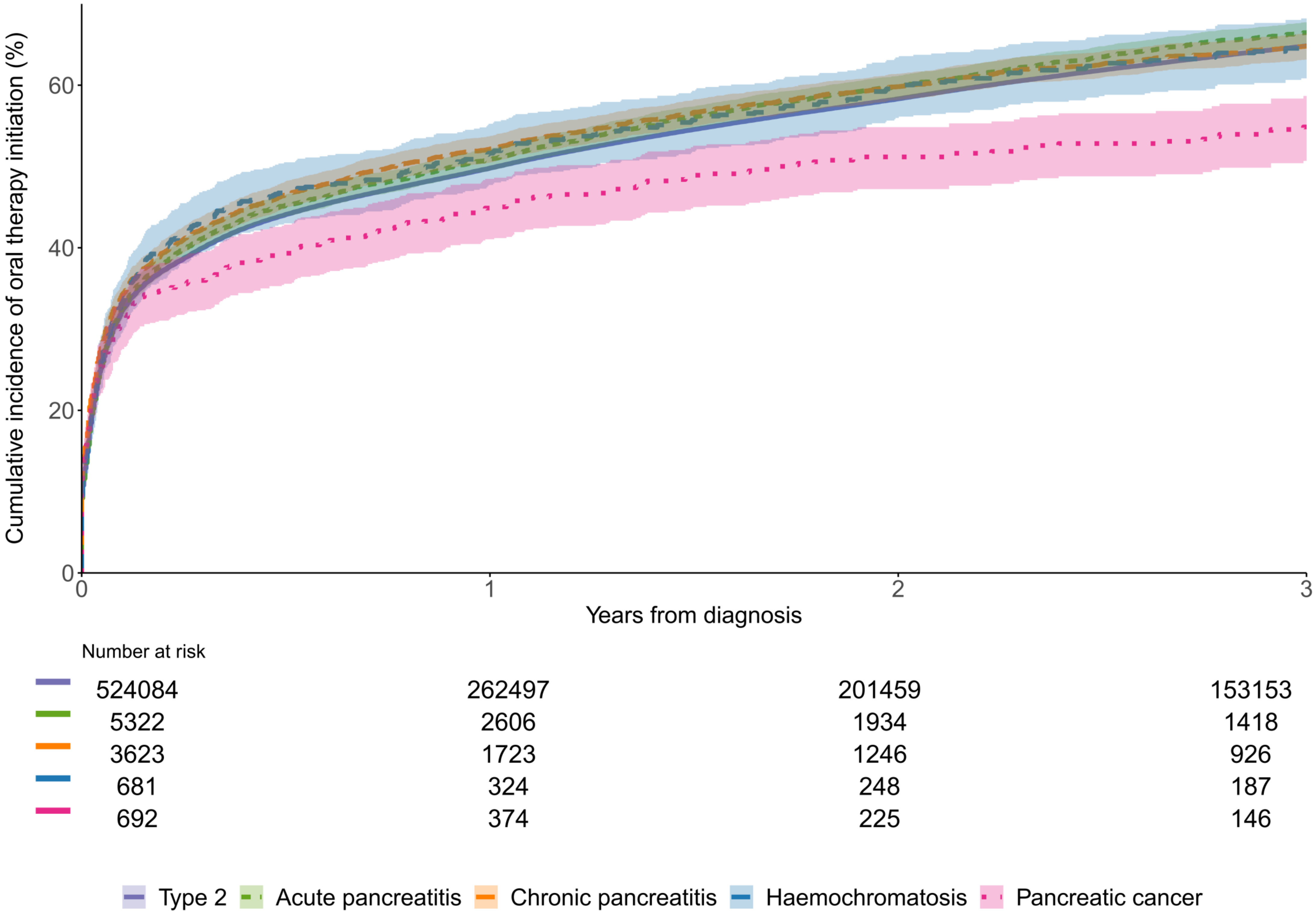

**Supplementary Table 1.** Baseline characteristics of matched cohort of individuals with type 3c diabetes and type 2 controls initiating a major glucose-lowering therapy class (metformin, sulphonylureas, thiazolidinediones [TZDs], SGLT2-inhibitors, DPP4-inhibitors).

| | **Metformin** | | **Sulphonylureas** | | **TZDs** | | **SGLT2-inhibitors** | | **DPP4-inhibitors** | |
|---|---|---|---|---|---|---|---|---|---|---|
| | **T2 controls** | **T3c** | **T2 controls** | **T3c** | **T2 controls** | **T3c** | **T2 controls** | **T3c** | **T2 controls** | **T3c** |
| n | 57310 | 6100 | 26056 | 3070 | 2947 | 381 | 7309 | 841 | 13745 | 1579 |
| **Sex** | | | | | | | | | | |
| Male | 32831 ( 57.3) | 3489 (57.2) | 15857 ( 60.9) | 1836 (59.8) | 1820 ( 61.8) | 226 (59.3) | 4131 ( 56.5) | 461 (54.8) | 7901 ( 57.5) | 887 (56.2) |
| Female | 24479 ( 42.7) | 2611 (42.8) | 10199 ( 39.1) | 1234 (40.2) | 1127 ( 38.2) | 155 (40.7) | 3178 ( 43.5) | 380 (45.2) | 5844 ( 42.5) | 692 (43.8) |
| **Age, years** | | | | | | | | | | |
| Median [IQR] | 61.42 [51.95, 70.68] | 61.19 [51.68, 70.71] | 61.63 [52.66, 70.98] | 60.57 [50.92, 70.42] | 58.70 [52.99, 65.80] | 59.03 [52.53, 66.89] | 58.79 [52.32, 66.11] | 58.67 [51.39, 66.46] | 63.20 [54.40, 71.76] | 63.17 [53.80, 72.04] |
| **Ethnicity** | | | | | | | | | | |
| White | 52799 ( 92.1) | 5510 (90.3) | 24890 ( 95.5) | 2836 (92.4) | 2877 ( 97.6) | 359 (94.2) | 6909 ( 94.5) | 763 (90.7) | 12929 ( 94.1) | 1437 (91.0) |
| South Asian | 2938 ( 5.1) | 342 ( 5.6) | 795 ( 3.1) | 150 ( 4.9) | 60 ( 2.0) | 17 ( 4.5) | 306 ( 4.2) | 51 ( 6.1) | 615 ( 4.5) | 98 ( 6.2) |
| Black | 1112 ( 1.9) | 143 ( 2.3) | 325 ( 1.2) | 62 ( 2.0) | 10 ( 0.3) | 5 ( 1.3) | 66 ( 0.9) | 13 ( 1.5) | 186 ( 1.4) | 36 ( 2.3) |
| Other | 148 ( 0.3) | 39 ( 0.6) | 7 ( 0.0) | 5 ( 0.2) | NA | NA | 4 ( 0.1) | 3 ( 0.4) | 3 ( 0.0) | 2 ( 0.1) |
| Mixed | 114 ( 0.2) | 37 ( 0.6) | 19 ( 0.1) | 9 ( 0.3) | NA | NA | 9 ( 0.1) | 7 ( 0.8) | 4 ( 0.0) | 4 ( 0.3) |
| Unknown | 199 ( 0.3) | 29 ( 0.5) | 20 ( 0.1) | 8 ( 0.3) | NA | NA | 15 ( 0.2) | 4 ( 0.5) | 8 ( 0.1) | 2 ( 0.1) |
| **Index of multiple deprivation quintile** | | | | | | | | | | |
| 1 (least deprived) | 9915 ( 17.3) | 1045 (17.1) | 4389 ( 16.8) | 499 (16.3) | 492 ( 16.7) | 66 (17.3) | 1306 ( 17.9) | 147 (17.5) | 2170 ( 15.8) | 261 (16.5) |
| 2 | 9701 ( 16.9) | 1025 (16.8) | 4487 ( 17.2) | 534 (17.4) | 445 ( 15.1) | 62 (16.3) | 1284 ( 17.6) | 149 (17.7) | 2293 ( 16.7) | 266 (16.8) |
| 3 | 10962 ( 19.1) | 1169 (19.2) | 5013 ( 19.2) | 590 (19.2) | 636 ( 21.6) | 76 (19.9) | 1427 ( 19.5) | 167 (19.9) | 2773 ( 20.2) | 314 (19.9) |
| 4 | 12246 ( 21.4) | 1304 (21.4) | 5623 ( 21.6) | 660 (21.5) | 575 ( 19.5) | 79 (20.7) | 1419 ( 19.4) | 163 (19.4) | 2880 ( 21.0) | 327 (20.7) |
| 5 (most deprived) | 14485 ( 25.3) | 1556 (25.5) | 6544 ( 25.1) | 787 (25.6) | 799 ( 27.1) | 98 (25.7) | 1873 ( 25.6) | 215 (25.6) | 3629 ( 26.4) | 411 (26.0) |
| **Alcohol consumption** | | | | | | | | | | |
| None | 3985 ( 7.3) | 498 ( 8.5) | 1769 ( 7.1) | 226 ( 7.7) | 249 ( 8.7) | 35 ( 9.4) | 428 ( 5.9) | 62 ( 7.4) | 938 ( 6.9) | 110 ( 7.0) |
| Within limits | 44332 ( 81.1) | 4067 (69.3) | 20244 ( 81.1) | 1939 (65.6) | 2280 ( 80.0) | 256 (68.4) | 6190 ( 85.3) | 624 (74.8) | 11359 ( 83.7) | 1147 (73.4) |
| Excess | 3841 ( 7.0) | 337 ( 5.7) | 1681 ( 6.7) | 174 ( 5.9) | 213 ( 7.5) | 22 ( 5.9) | 314 ( 4.3) | 40 ( 4.8) | 726 ( 5.3) | 72 ( 4.6) |
| Harmful | 2484 ( 4.5) | 970 (16.5) | 1257 ( 5.0) | 615 (20.8) | 108 ( 3.8) | 61 (16.3) | 322 ( 4.4) | 108 (12.9) | 552 ( 4.1) | 234 (15.0) |
| Unknown | 2668 ( 4.7) | 228 ( 3.7) | 1105 ( 4.2) | 116 ( 3.8) | 97 ( 3.3) | 7 ( 1.8) | 55 ( 0.8) | 7 ( 0.8) | 170 ( 1.2) | 16 ( 1.0) |
| **HbA1c, mmol/mol** | | | | | | | | | | |
| Median [IQR] | 61.87 [54.00, 78.14] | 62.83 [54.00, 82.00] | 74.00 [62.83, 94.33] | 75.00 [62.00, 95.00] | 71.58 [62.92, 84.00] | 72.84 [62.92, 85.79] | 73.00 [64.00, 86.00] | 73.00 [64.00, 85.00] | 68.00 [61.00, 80.00] | 69.00 [60.00, 81.00] |
| Missing | 8419 (14.7) | 1064 (17.4) | 4940 (19.0) | 701 (22.8) | 428 (14.5) | 76 (19.9) | 790 (10.8) | 120 (14.3) | 1439 (10.5) | 212 (13.4) |
| **BMI, kg/m2** | | | | | | | | | | |
| Median [IQR] | 32.10 [28.30, 36.90] | 30.60 [26.50, 35.50] | 31.00 [27.23, 35.50] | 28.60 [24.70, 33.30] | 32.10 [28.70, 36.40] | 29.45 [25.60, 34.69] | 33.50 [29.77, 38.20] | 32.17 [27.80, 37.30] | 32.00 [28.22, 36.50] | 30.40 [26.20, 35.00] |
| Missing | 8472 (14.8) | 981 (16.1) | 2825 (10.8) | 407 (13.3) | 86 (2.9) | 13 (3.4) | 275 (3.8) | 29 (3.4) | 698 (5.1) | 73 (4.6) |
| **Weight, kg** | | | | | | | | | | |
| Median [IQR] | 92.00 [79.50, 107.00] | 87.00 [74.00, 102.00] | 89.20 [76.60, 104.00] | 81.50 [69.85, 96.00] | 93.00 [80.90, 107.00] | 83.85 [70.11, 101.75] | 97.00 [84.20, 112.00] | 92.10 [79.00, 108.00] | 91.80 [79.40, 106.00] | 86.00 [74.00, 101.00] |
| Missing | 6764 (11.8) | 760 (12.5) | 2192 (8.4) | 315 (10.3) | 54 (1.8) | 7 (1.8) | 159 (2.2) | 15 (1.8) | 461 (3.4) | 58 (3.7) |
| **Number of other glucose-lowering therapies being taken** | | | | | | | | | | |

| | Metformin | | Sulphonylureas | | TZDs | | SGLT2-inhibitors | | DPP4-inhibitors | |
|---|---|---|---|---|---|---|---|---|---|---|
| | **T2 controls** | **T3c** | **T2 controls** | **T3c** | **T2 controls** | **T3c** | **T2 controls** | **T3c** | **T2 controls** | **T3c** |
| 0 | 53940 ( 94.1) | 5530 (90.7) | 8742 ( 33.6) | 1271 (41.4) | 201 ( 6.8) | 43 (11.3) | 558 ( 7.6) | 81 ( 9.6) | 2018 ( 14.7) | 300 (19.0) |
| 1 | 3275 ( 5.7) | 556 ( 9.1) | 15371 ( 59.0) | 1628 (53.0) | 1566 ( 53.1) | 196 (51.4) | 3418 ( 46.8) | 415 (49.3) | 7913 ( 57.6) | 865 (54.8) |
| 2+ | 95 ( 0.2) | 14 ( 0.2) | 1943 ( 7.5) | 171 ( 5.6) | 1180 ( 40.0) | 142 (37.3) | 3333 ( 45.6) | 345 (41.0) | 3814 ( 27.7) | 414 (26.2) |
| **PEI status** | | | | | | | | | | |
| No PEI | NA | 5250 (86.1) | NA | 2435 (79.3) | NA | 325 (85.3) | NA | 724 (86.1) | NA | 1384 (87.7) |
| PEI | NA | 850 (13.9) | NA | 635 (20.7) | NA | 56 (14.7) | NA | 117 (13.9) | NA | 195 (12.3) |
| **3c subtype** | | | | | | | | | | |
| Acute pancreatitis only | NA | 3516 (57.6) | NA | 1585 (51.6) | NA | 207 (54.3) | NA | 515 (61.2) | NA | 943 (59.7) |
| Chronic pancreatitis | NA | 1882 (30.9) | NA | 1143 (37.2) | NA | 147 (38.6) | NA | 239 (28.4) | NA | 469 (29.7) |
| Haemochromatosis | NA | 452 ( 7.4) | NA | 171 ( 5.6) | NA | 15 ( 3.9) | NA | 66 ( 7.8) | NA | 115 ( 7.3) |
| Pancreatic cancer | NA | 250 ( 4.1) | NA | 171 ( 5.6) | NA | 12 ( 3.1) | NA | 21 ( 2.5) | NA | 52 ( 3.3) |

**Supplementary Figure 7.** A) Mean HbA1c response and B) Proportion of early treatment discontinuation, in individuals with type 3c diabetes following pancreatic cancer with PEI (orange) and without PEI (green), and matched type 2 controls (blue/purple) initiating an oral glucose lowering therapy. Contrasts represent estimated differences between groups with 95% confidence intervals for HbA1c change in mmol/mol and odds ratios with 95% confidence intervals for discontinuation. Models were adjusted for baseline HbA1c and number of other glucose-lowering therapies being taken.

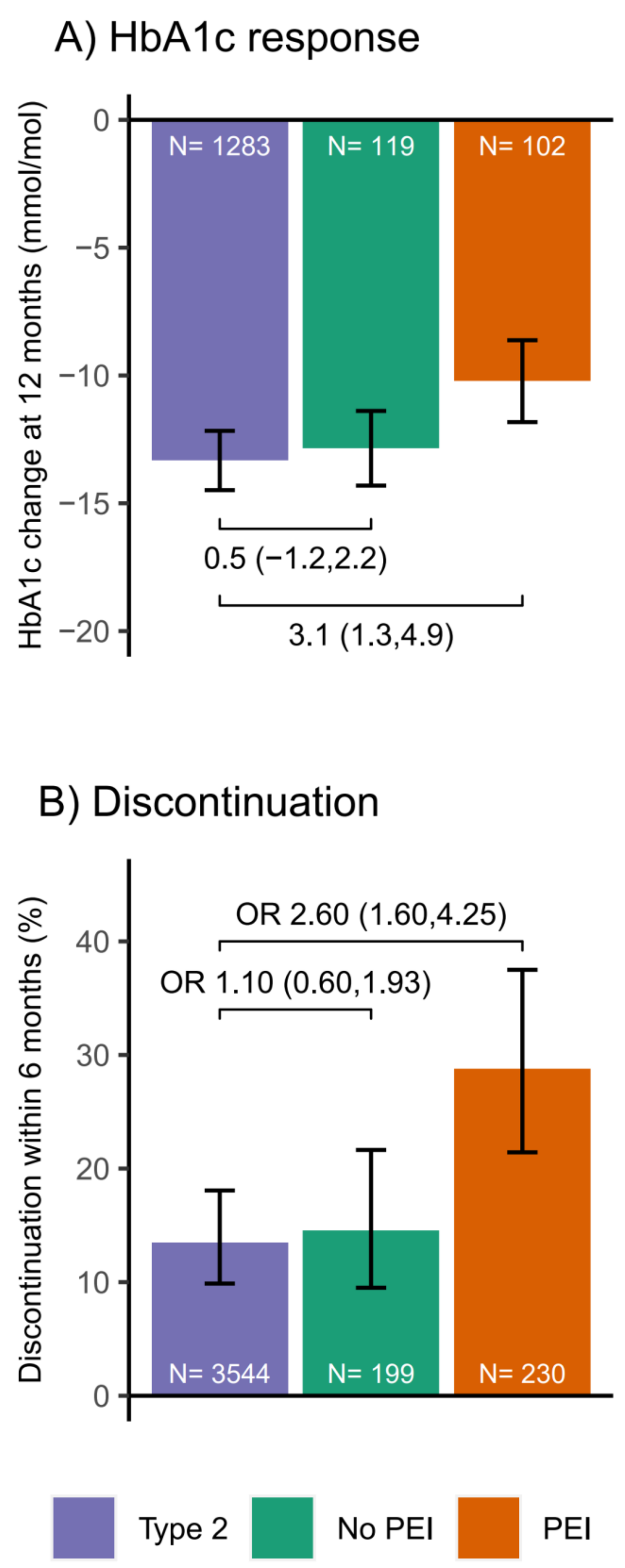

**Supplementary Figure 8.** A) Mean HbA1c response and B) Proportion of early treatment discontinuation, in individuals with type 3c diabetes following haemochromatosis without PEI (green), and matched type 2 controls (blue/purple) initiating an oral glucose lowering therapy. Contrasts represent estimated differences between groups with 95% confidence intervals for HbA1c change in mmol/mol and odds ratios with 95% confidence intervals for discontinuation. Models were adjusted for baseline HbA1c and number of other glucose-lowering therapies being taken.

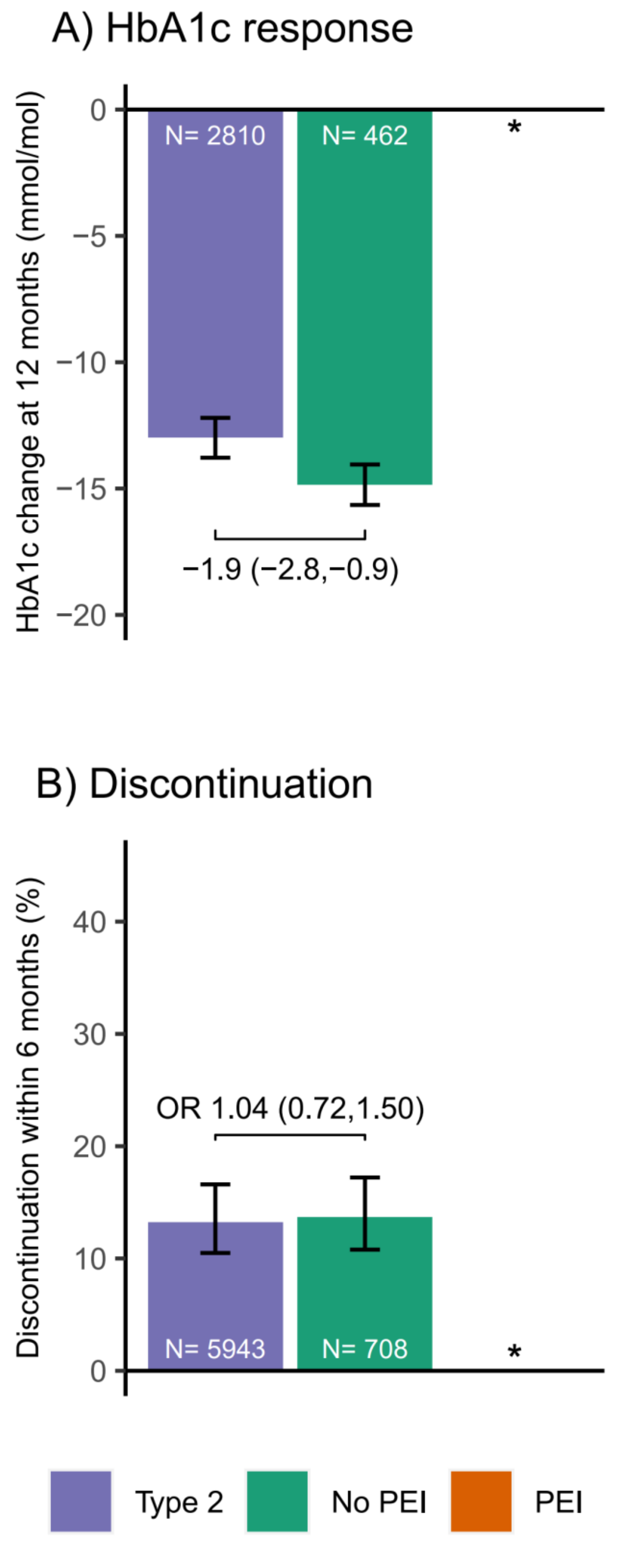


*Due to small numbers of individuals with type 3c diabetes following haemochromatosis with PEI these individuals were not included in this analysis.

**Supplementary table 2.** Treatment response outcomes by drug class in all individuals with type 3c diabetes and type 2 controls.

**A) HbA1c**

| Drug class | Type 2 | | Type 3c PEI | | | Type 3c No PEI | | |
|---|---|---|---|---|---|---|---|---|
| | N | Mean HbA1c change (95% CI) | N | Mean HbA1c change (95% CI) | Mean difference vs Type 2 (95% CI) | N | Mean HbA1c change (95% CI) | Mean difference vs Type 2 (95% CI) |
| Metformin | 22581 | -17.4 (-17.6,-17.1) | 404 | -13.1 (-13.8,-12.4) | 4.2 (3.5,5.0) | 3090 | -17.2 (-17.4,-16.9) | 0.2 (-0.1,0.5) |
| Sulphonylureas | 6398 | -14.2 (-14.8,-13.7) | 226 | -10.1 (-11.4,-8.8) | 4.2 (2.8,5.5) | 1095 | -12.6 (-13.2,-12.0) | 1.6 (0.9,2.4) |
| TZDs | 800 | -12.9 (-14.4,-11.5) | 18 | -10.7 (-14.6,-6.7) | 2.3 (-1.9,6.4) | 139 | -12.6 (-14.1,-11.1) | 0.3 (-1.6,2.3) |
| DPP4–inhibitors | 4634 | -8.2 (-8.8,-7.6) | 83 | -6.9 (-8.7,-5.1) | 1.3 (-0.6,3.2) | 743 | -7.4 (-8.0,-6.8) | 0.8 (0,1.7.0) |
| SGLT2–inhibitors | 1871 | -11.5 (-12.4,-10.7) | 51 | -12.9 (-15.2,-10.7) | -1.4 (-3.8,1.0) | 341 | -10.5 (-11.4,-9.5) | 1.1 (-0.1,2.3) |

**B) Discontinuation**

| Drug class | Type 2 | | Type 3c PEI | | | Type 3c No PEI | | |
|---|---|---|---|---|---|---|---|---|
| | N | Mean discontinuation (95% CI) | N | Mean discontinuation (95% CI) | Odds ratio vs Type 2 (95% CI) | N | Mean discontinuation (95% CI) | Odds ratio vs Type 2 (95% CI) |
| Metformin | 45194 | 5.1 (4.5,5.8) | 726 | 13.9 (11.3,16.8) | 2.97 (2.29,3.84) | 4638 | 6.4 (5.6,7.2) | 1.26 (1.05,1.50) |
| Sulphonylureas | 21084 | 17.8 (16.1,19.5) | 546 | 24.6 (20.5,29.2) | 1.51 (1.17,1.94) | 2148 | 17.7 (15.8,19.7) | 0.99 (0.84,1.17) |
| TZDs | 2536 | 19.2 (14.8,24.5) | 47 | 24.6 (13.1,41.2) | 1.37 (0.58,2.99) | 299 | 19.8 (15.0,25.7) | 1.04 (0.68,1.58) |
| DPP4–inhibitors | 11198 | 16.0 (14.0,18.2) | 165 | 23.8 (17.5,31.6) | 1.64 (1.07,2.47) | 1214 | 17.8 (15.6,20.2) | 1.14 (0.91,1.41) |
| SGLT2–inhibitors | 5580 | 21.4 (18.1,25.2) | 94 | 27.8 (1.09,38.6) | 1.41 (0.81,2.36) | 613 | 20.6 (17.0,24.6) | 0.95 (0.71,1.27) |

**C) Weight**

| Drug class | Type 2 | | Type 3c PEI | | | Type 3c No PEI | | |
|---|---|---|---|---|---|---|---|---|
| | N | Mean weight change (95% CI) | N | Mean weight change (95% CI) | Mean difference vs Type 2 (95% CI) | N | Mean weight change (95% CI) | Mean difference vs Type 2 (95% CI) |
| Metformin | 19770 | -2.7 (-2.8,-2.6) | 396 | -3.2 (-3.5,-2.8) | -0.4 (-0.8,-0.1) | 2934 | -2.9 (-3.0,-2.7) | -0.1 (-0.3,0.0) |
| Sulphonylureas | 7374 | 1.3 (1.2,1.5) | 267 | 0.2 (-0.2,0.6) | -1.1 (-1.5,-0.7) | 1191 | 1.1 (0.9,1.3) | -0.2 (-0.4,0.0) |
| TZDs | 1031 | 2.0 (1.5,2.5) | 22 | 1.0 (-0.4,2.5) | -1.0 (-2.5,0.5) | 171 | 1.7 (1.2,2.2) | -0.3 (-1.0,0.4) |
| DPP4–inhibitors | 4297 | -1.3 (-1.5,-1.1) | 88 | -1.5 (-2.1,-0.9) | -0.2 (-0.8,0.5) | 703 | -0.9 (-1.1,-0.7) | 0.4 (0.1,0.7) |
| SGLT2–inhibitors | 1895 | -3.8 (-4.2,-3.5) | 54 | -5.3 (-6.1,-4.4) | -1.4 (-2.3,-0.6) | 351 | -3.9 (-4.2,-3.5) | 0.0 (-0.5,0.4) |

**Supplementary table 3.** Treatment response outcomes by drug class in individuals with type 3c diabetes following acute pancreatitis and type 2 controls.

**A) HbA1c**

| Drug class | Type 2 | | Type 3c PEI | | | Type 3c No PEI | | |
|---|---|---|---|---|---|---|---|---|
| | **N** | **Mean HbA1c change (95% CI)** | **N** | **Mean HbA1c change (95% CI)** | **Mean difference vs Type 2 (95% CI)** | **N** | **Mean HbA1c change (95% CI)** | **Mean difference vs Type 2 (95% CI)** |
| Metformin | 13667 | -17.5 (-17.8,-17.2) | 51 | -15.3 (-17.1,-13.4) | 2.3 (0.4,4.2) | 2053 | -17.4 (-17.7,-17.0) | 0.2 (-0.2,0.6) |
| Sulphonylureas | 3673 | -14.2 (-14.8,-13.5) | 29 | -10.3 (-13.5,-7.0) | 3.9 (0.6,7.2) | 713 | -13.0 (-13.7,-12.3) | 1.1 (0.2,2.1) |
| TZDs* | 445 | -13.2 (-15.3,-11.1) | | | | 88 | -13.0 (-15.0,-11.0) | 0.2 (-2.5,2.9) |
| DPP4–inhibitors | 2878 | -8.5 (-9.2,-7.7) | 18 | -5.6 (-9.3,-1.8) | 2.9 (-1.0,6.7) | 494 | -6.8 (-7.5,-6.1) | 1.7 (0.6,2.7) |
| SGLT2–inhibitors | 1166 | -11.0 (-12.2,-9.9) | 17 | -16.5 (-20.5,-12.4) | -5.5 (-9.6,-1.3) | 229 | -9.8 (-11.0,-8.6) | 1.2 (-0.3,2.7) |

**B) Discontinuation**

| Drug class | Type 2 | | Type 3c PEI | | | Type 3c No PEI | | |
|---|---|---|---|---|---|---|---|---|
| | **N** | **Mean discontinuation (95% CI)** | **N** | **Mean discontinuation (95% CI)** | **Odds ratio vs Type 2 (95% CI)** | **N** | **Mean discontinuation (95% CI)** | **Odds ratio vs Type 2 (95% CI)** |
| Metformin | 26180 | 5.2 (4.4,6.1) | 85 | 7.6 (3.4,16.1) | 1.49 (0.56,3.27) | 3009 | 5.8 (4.9,6.8) | 1.12 (0.89,1.42) |
| Sulphonylureas | 11016 | 16.9 (14.8,19.3) | 57 | 20.4 (10.9,34.8) | 1.26 (0.56,2.55) | 1341 | 17.0 (14.8,19.5) | 1.01 (0.81,1.26) |
| TZDs* | 1390 | 18.5 (13.0,25.7) | | | | 187 | 19.4 (13.7,26.7) | 1.06 (0.61,1.85) |
| DPP4–inhibitors | 6589 | 15.7 (13.2,18.5) | 30 | 10.8 (3.5,28.8) | 0.65 (0.15,1.91) | 786 | 18.4 (15.6,21.4) | 1.21 (0.92,1.59) |
| SGLT2–inhibitors | 3450 | 21.3 (17.2,26.1) | 26 | 16.3 (6.2,36.4) | 0.72 (0.21,1.96) | 417 | 18.4 (14.4,23.3) | 0.83 (0.58,1.20) |

**C) Weight**

| Drug class | Type 2 | | Type 3c PEI | | | Type 3c No PEI | | |
|---|---|---|---|---|---|---|---|---|
| | **N** | **Mean weight change (95% CI)** | **N** | **Mean weight change (95% CI)** | **Mean difference vs Type 2 (95% CI)** | **N** | **Mean weight change (95% CI)** | **Mean difference vs Type 2 (95% CI)** |
| Metformin | 11944 | -2.9 (-3.0,-2.7) | 49 | -3.2 (-4.1,-2.3) | -0.3 (-1.2,0.6) | 1956 | -2.8 (-2.9,-2.6) | 0.1 (-0.1,0.3) |
| Sulphonylureas | 4115 | 1.2 (0.9,1.4) | 36 | -0.2 (-1.2,0.8) | -1.4 (-2.4,-0.4) | 760 | 1.3 (1.1,1.6) | 0.2 (-0.1,0.5) |
| TZDs* | 598 | 1.7 (1,2.4.0) | | | | 109 | 1.2 (0.5,2.0) | -0.5 (-1.4,0.5) |
| DPP4–inhibitors | 2621 | -1.3 (-1.5,-1.0) | 20 | -1.4 (-2.7,-0.1) | -0.1 (-1.4,1.2) | 453 | -0.8 (-1.1,-0.6) | 0.4 (0.1,0.8) |
| SGLT2–inhibitors | 1208 | -3.9 (-4.3,-3.4) | 15 | -5.1 (-6.6,-3.5) | -1.2 (-2.8,0.4) | 245 | -4.0 (-4.4,-3.6) | -0.2 (-0.7,0.4) |

*No results shown for T3c PEI where N <10 for this group.

**Supplementary table 4.** Treatment response outcomes by drug class in individuals with type 3c diabetes following chronic pancreatitis and type 2 controls.

**A) HbA1c**

| Drug class | Type 2 | | Type 3c PEI | | | Type 3c No PEI | | |
|---|---|---|---|---|---|---|---|---|
| | N | Mean HbA1c change (95% CI) | N | Mean HbA1c change (95% CI) | Mean difference vs Type 2 (95% CI) | N | Mean HbA1c change (95% CI) | Mean difference vs Type 2 (95% CI) |
| Metformin | 6297 | -16.9 (-17.4,-16.4) | 304 | -12.7 (-13.6,-11.8) | 4.2 (3.2,5.2) | 684 | -15.6 (-16.2,-15.0) | 1.3 (0.5,2.1) |
| Sulphonylureas | 2028 | -14.2 (-15.2,-13.1) | 158 | -9.5 (-11.2,-7.7) | 4.7 (2.8,6.7) | 279 | -10.6 (-11.9,-9.3) | 3.6 (2,5.2.0) |
| TZDs | 289 | -12.1 (-14.3,-10.0) | 15 | -9.9 (-13.8,-5.9) | 2.2 (-2.1,6.6) | 41 | -11.5 (-14.2,-8.9) | 0.6 (-2.6,3.8) |
| DPP4–inhibitors | 1209 | -8.0 (-9.2,-6.8) | 55 | -7.2 (-9.5,-4.9) | 0.7 (-1.8,3.3) | 167 | -8.0 (-9.4,-6.7) | -0.1 (-1.8,1.7) |
| SGLT2–inhibitors | 529 | -12.8 (-14.4,-11.3) | 29 | -11.7 (-14.5,-8.9) | 1.2 (-1.9,4.3) | 79 | -10.8 (-12.5,-9.0) | 2.1 (-0.1,4.2) |

**B) Discontinuation**

| Drug class | Type 2 | | Type 3c PEI | | | Type 3c No PEI | | |
|---|---|---|---|---|---|---|---|---|
| | N | Mean discontinuation (95% CI) | N | Mean discontinuation (95% CI) | Odds ratio vs Type 2 (95% CI) | N | Mean discontinuation (95% CI) | Odds ratio vs Type 2 (95% CI) |
| Metformin | 13831 | 5.0 (4.0,6.3) | 525 | 13.9 (11.0,17.4) | 3.06 (2.15,4.35) | 1135 | 8.3 (6.7,10.3) | 1.72 (1.24,2.38) |
| Sulphonylureas | 7804 | 18.8 (16.1,21.9) | 406 | 25.3 (20.4,30.8) | 1.46 (1.06,1.99) | 608 | 18.8 (15.3,22.9) | 1.00 (0.74,1.34) |
| TZDs | 938 | 21.6 (14.3,31.3) | 39 | 28.5 (14.8,47.8) | 1.44 (0.54,3.64) | 90 | 19.1 (10.9,31.3) | 0.86 (0.40,1.80) |
| DPP4–inhibitors | 3336 | 16.1 (12.6,20.4) | 113 | 25.0 (17.4,34.5) | 1.74 (1.01,2.93) | 299 | 18.5 (14.1,23.9) | 1.18 (0.77,1.80) |
| SGLT2–inhibitors | 1543 | 21.6 (15.6,29.2) | 60 | 32.1 (20.3,46.7) | 1.72 (0.82,3.49) | 133 | 21.0 (14.0,30.1) | 0.96 (0.53,1.72) |

**C) Weight**

| Drug class | Type 2 | | Type 3c PEI | | | Type 3c No PEI | | |
|---|---|---|---|---|---|---|---|---|
| | N | Mean weight change (95% CI) | N | Mean weight change (95% CI) | Mean difference vs Type 2 (95% CI) | N | Mean weight change (95% CI) | Mean difference vs Type 2 (95% CI) |
| Metformin | 5476 | -2.5 (-2.7,-2.3) | 292 | -3.0 (-3.4,-2.6) | -0.5 (-0.9,0.0) | 649 | -3.2 (-3.4,-2.9) | -0.6 (-1.0,-0.3) |
| Sulphonylureas | 2493 | 1.5 (1.3,1.8) | 195 | 0.2 (-0.3,0.8) | -1.3 (-1.9,-0.7) | 312 | 0.7 (0.3,1.0) | -0.9 (-1.3,-0.4) |
| TZDs | 354 | 2.7 (1.9,3.4) | 18 | 1.6 (0.2,3.0) | -1.1 (-2.6,0.5) | 50 | 3.0 (2.1,3.9) | 0.3 (-0.8,1.4) |
| DPP4–inhibitors | 1194 | -1.5 (-1.8,-1.1) | 56 | -1.4 (-2.1,-0.7) | 0.0 (-0.7,0.8) | 174 | -0.9 (-1.3,-0.5) | 0.6 (0,1.1.0) |
| SGLT2–inhibitors | 517 | -4.0 (-4.7,-3.3) | 37 | -4.9 (-5.9,-3.9) | -0.9 (-2.0,0.3) | 72 | -3.2 (-3.9,-2.5) | 0.8 (-0.1,1.7) |

**Supplementary table 5.** Treatment response outcomes by drug class in individuals with type 3c diabetes following pancreatic cancer and type 2 controls.

**A) HbA1c**

| **Drug class** | **Type 2** | | **Type 3c PEI** | | | **Type 3c No PEI** | | |
|---|---|---|---|---|---|---|---|---|
| | **N** | **Mean HbA1c change (95% CI)** | **N** | **Mean HbA1c change (95% CI)** | **Mean difference vs Type 2 (95% CI)** | **N** | **Mean HbA1c change (95% CI)** | **Mean difference vs Type 2 (95% CI)** |
| Metformin | 754 | -18.1 (-19.4,-16.9) | 49 | -14.9 (-16.8,-12.9) | 3.2 (1,5.5) | 68 | -17.3 (-18.9,-15.7) | 0.8 (-1.2,2.9) |
| Sulphonylureas | 302 | -14.2 (-16.9,-11.6) | 38 | -9 (-12.6,-5.5) | 5.2 (1.4,9) | 26 | -13.4 (-17.2,-9.5) | 0.9 (-3.4,5.1) |
| TZDs* | | | | | | | | |
| DPP4–inhibitors** | 172 | -6.4 (-9.2,-3.6) | | | | 18 | -10.6 (-14.0,-7.2) | -4.2 (-8.5,0.1) |
| SGLT2–inhibitors* | | | | | | | | |

**B) Discontinuation**

| **Drug class** | **Type 2** | | **Type 3c PEI** | | | **Type 3c No PEI** | | |
|---|---|---|---|---|---|---|---|---|
| | **N** | **Mean discontinuation (95% CI)** | **N** | **Mean discontinuation (95% CI)** | **Odds ratio vs Type 2 (95% CI)** | **N** | **Mean discontinuation (95% CI)** | **Odds ratio vs Type 2 (95% CI)** |
| Metformin | 1808 | 5.7 (3.1,10.1) | 115 | 19.7 (12.4,29.9) | 4.09 (1.86,9.38) | 99 | 12.3 (6.7,21.3) | 2.33 (0.95,5.72) |
| Sulphonylureas | 1120 | 19.9 (12.4,30.5) | 82 | 28.3 (16.1,44.7) | 1.59 (0.69,3.59) | 56 | 8.0 (2.6,22.5) | 0.35 (0.08,1.14) |
| TZDs* | | | | | | | | |
| DPP4–inhibitors | 394 | 13.3 (6.0,27.0) | 21 | 40.2 (18.8,66.1) | 4.38 (1.11,18.06) | 27 | 4.7 (0.6,26.9) | 0.32 (0.02,2.07) |
| SGLT2–inhibitors** | 150 | 20.8 (6.5,50.0) | | | | 10 | 49.0 (15.8,83.1) | 3.65 (0.47,33.34) |

**C) Weight**

| **Drug class** | **Type 2** | | **Type 3c PEI** | | | **Type 3c No PEI** | | |
|---|---|---|---|---|---|---|---|---|
| | **N** | **Mean weight change (95% CI)** | **N** | **Mean weight change (95% CI)** | **Mean difference vs Type 2 (95% CI)** | **N** | **Mean weight change (95% CI)** | **Mean difference vs Type 2 (95% CI)** |
| Metformin | 696 | -3.0 (-3.6,-2.5) | 54 | -4.7 (-5.5,-3.9) | -1.7 (-2.6,-0.7) | 59 | -5.3 (-6.1,-4.6) | -2.3 (-3.2,-1.4) |
| Sulphonylureas | 324 | 1.6 (0.6,2.5) | 36 | -0.6 (-2.1,0.8) | -2.2 (-3.7,-0.6) | 30 | 1.0 (-0.5,2.5) | -0.5 (-2.2,1.1) |
| TZDs* | | | | | | | | |
| DPP4–inhibitors | 186 | -1.2 (-2.2,-0.2) | 11 | -1.5 (-3.3,0.2) | -0.3 (-2.3,1.7) | 20 | -0.1 (-1.3,1.2) | 1.1 (-0.5,2.7) |
| SGLT2–inhibitors* | | | | | | | | |

*No results shown for drug classes where T3c PEI and T3c No PEI groups both have N <10.

**No results shown for T3c PEI where N <10 for this group.

**Supplementary table 6.** Treatment response outcomes by drug class in individuals with type 3c diabetes following haemochromatosis and type 2 controls.

**A) HbA1c**

| **Drug class** | **Type 2** | | **Type 3c No PEI** | | |
|---|---|---|---|---|---|
| | **N** | **Mean HbA1c change (95% CI)** | **N** | **Mean HbA1c change (95% CI)** | **Mean difference vs Type 2 (95% CI)** |
| Metformin | 1863 | -17.4 (-18.2,-16.6) | 285 | -19.2 (-20.0,-18.4) | -1.8 (-2.8,-0.7) |
| Sulphonylureas | 390 | -13.5 (-15.6,-11.3) | 77 | -15.9 (-18.1,-13.7) | -2.5 (-5.3,0.4) |
| TZDs* | | | | | |
| DPP4–inhibitors | 370 | -8.2 (-10.5,-5.9) | 64 | -9.4 (-11.7,-7.2) | -1.3 (-4.4,1.8) |
| SGLT2–inhibitors | 136 | -9.7 (-12.7,-6.7) | 28 | -13.7 (-16.9,-10.5) | -4.0 (-7.8,-0.2) |

**B) Discontinuation**

| **Drug class** | **Type 2** | | **Type 3c No PEI** | | |
|---|---|---|---|---|---|
| | **N** | **Mean discontinuation (95% CI)** | **N** | **Mean discontinuation (95% CI)** | **Odds ratio vs Type 2 (95% CI)** |
| Metformin | 3367 | 4.7 (2.9,7.7) | 395 | 3.5 (2.0,6.2) | 0.73 (0.34,1.55) |
| Sulphonylureas | 1134 | 19.6 (12.9,28.6) | 143 | 22.8 (15.3,32.6) | 1.21 (0.64,2.31) |
| TZDs | 136 | 8.1 (1.2,39.8) | 15 | 18.9 (4.2,55.2) | 2.65 (0.29,35.65) |
| DPP4–inhibitors | 869 | 18.9 (12.0,28.6) | 102 | 13.4 (7.8,22.2) | 0.66 (0.29,1.49) |
| SGLT2–inhibitors | 437 | 21.0 (10.6,37.2) | 53 | 32.1 (18.4,49.8) | 1.78 (0.68,4.91) |

**C) Weight**

| **Drug class** | **Type 2** | | **Type 3c No PEI** | | |
|---|---|---|---|---|---|
| | **N** | **Mean weight change (95% CI)** | **N** | **Mean weight change (95% CI)** | **Mean difference vs Type 2 (95% CI)** |
| Metformin | 1648 | -2.3 (-2.7,-1.9) | 270 | -2.5 (-2.9,-2.1) | -0.2 (-0.7,0.4) |
| Sulphonylureas | 442 | 1.9 (1.0,2.9) | 89 | 1.0 (0.0,1.9) | -1.0 (-2.2,0.2) |
| TZDs* | | | | | |
| DPP4–inhibitors | 290 | -1.1 (-1.9,-0.2) | 56 | -1.4 (-2.2,-0.5) | -0.3 (-1.4,0.9) |
| SGLT2–inhibitors | 139 | -3.4 (-4.8,-1.9) | 29 | -4.0 (-5.6,-2.5) | -0.7 (-2.4,1.1) |

*No results shown for T3c No PEI group where N <10 for this group.

**Supplementary Figure 9.** Sensitivity analysis restricted to individuals whose alcohol consumption was within recommended limits or who did not consume alcohol.

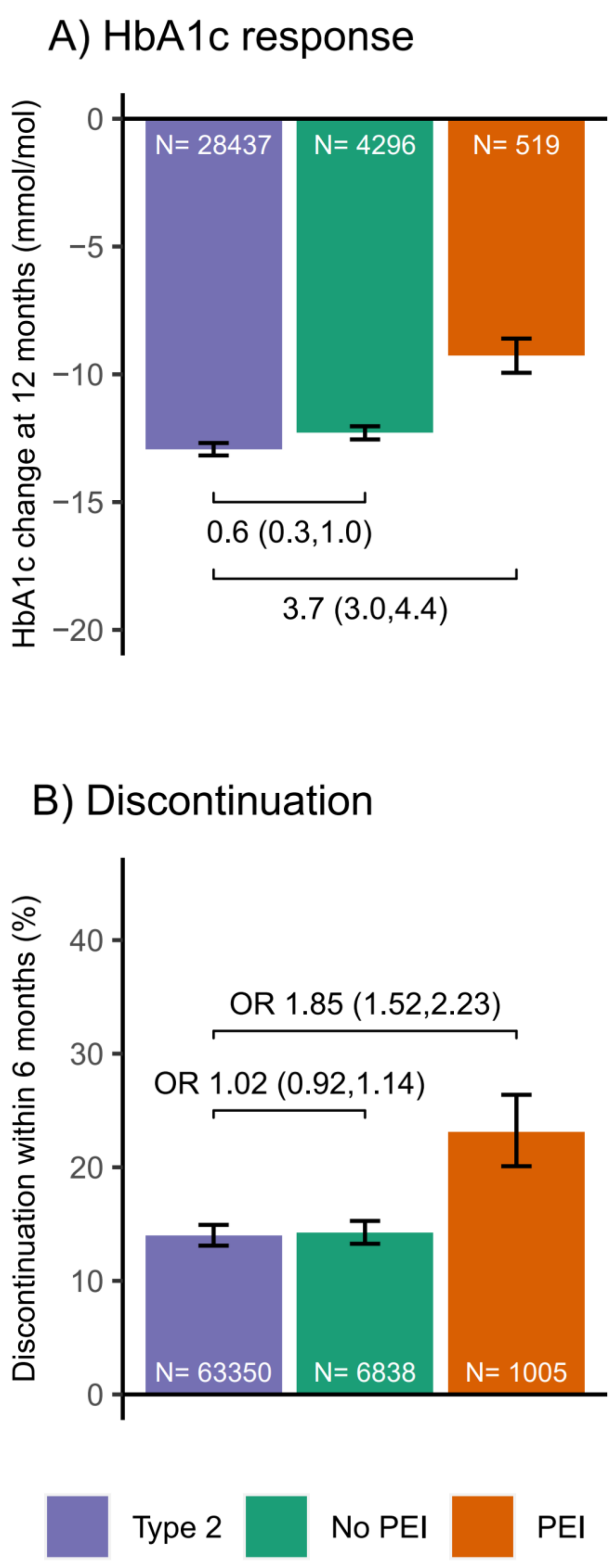

**Supplementary Figure 10.** Sensitivity analysis restricting the criteria of the PEI subgroup to those with a PERT prescription within the 6 months prior to drug initiation.

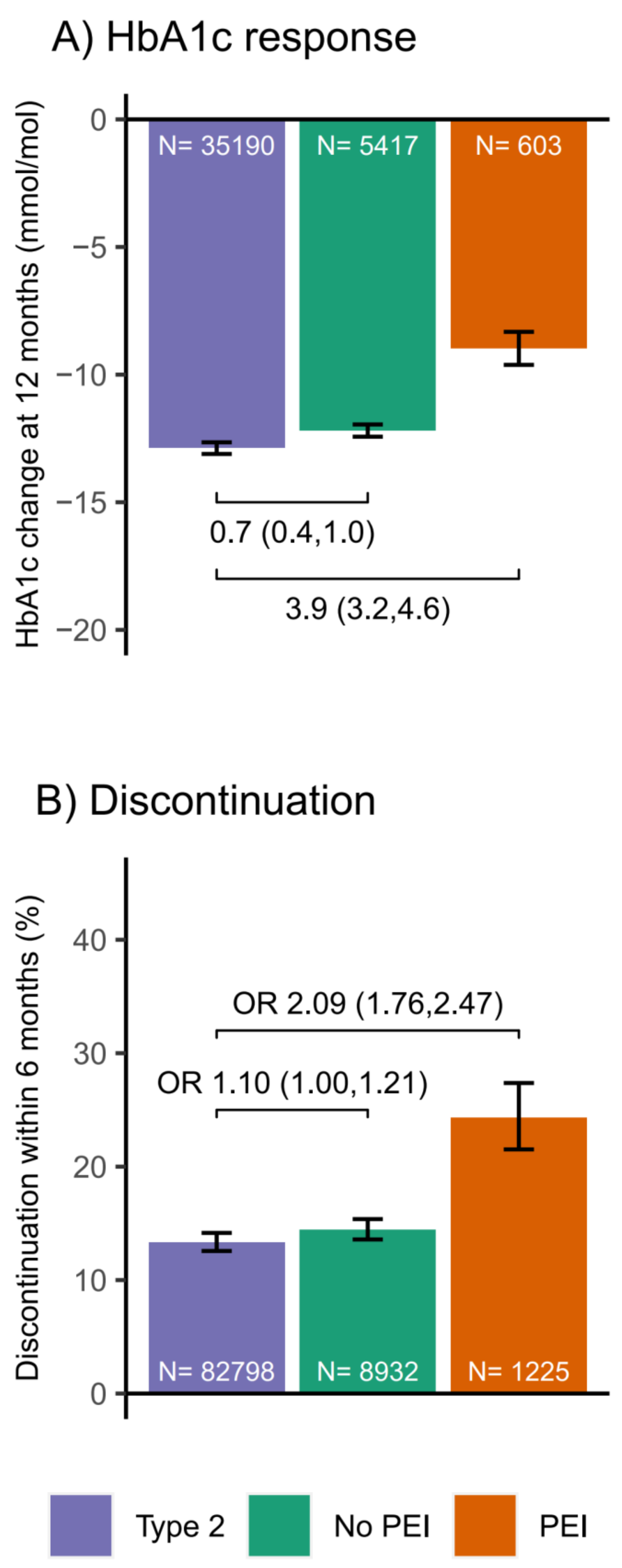

**Supplementary Figure 11.** Sensitivity analysis restricting the criteria of diabetes following acute pancreatitis to those with a record of acute pancreatitis within the 5 years prior to diabetes diagnosis.

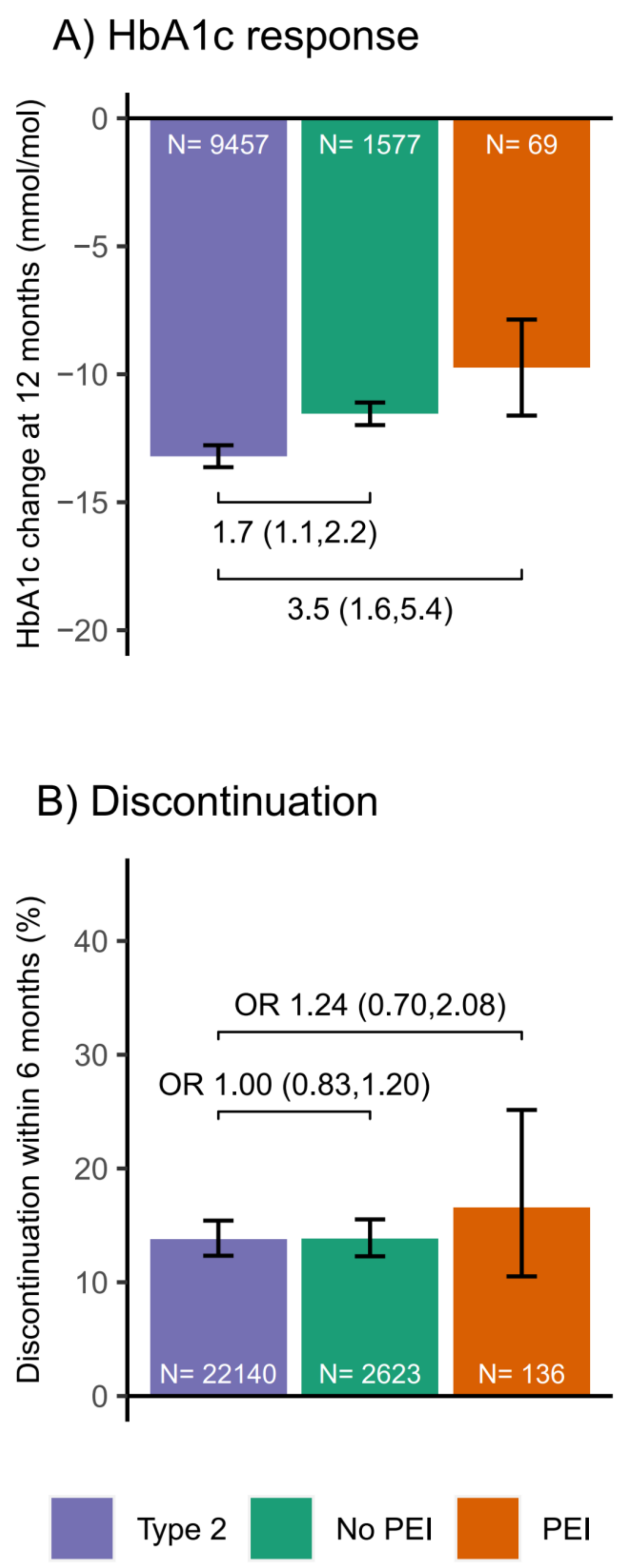
